# Risk of cardiovascular disease after a diagnosis of common psychiatric disorders: a matched cohort study of disease susceptibility and progression trajectory in the UK Biobank

**DOI:** 10.1101/2021.09.28.21264283

**Authors:** Xin Han, Yu Zeng, Yannan Shang, Yao Hu, Can Hou, Huazhen Yang, Wenwen Chen, Zhiye Ying, Yajing Sun, Yuanyuan Qu, Junren Wang, Wei Zhang, Fang Fang, Unnur A. Valdimarsdóttir, Huan Song

## Abstract

**Background:** Whether associations between psychiatric disorders and cardiovascular diseases (CVDs) can be modified by disease susceptibility and the temporal pattern of these associated CVDs remain unknown.

**Methods:** We conducted a matched cohort study of UK Biobank including 35,227 patients with common psychiatry disorders (anxiety, depression, and stress-related disorders) between 1997 and 2019, together with 176,135 sex- and birth year-individually matched unexposed individuals.

**Results:** The mean age at the index date was 51.76 years, and 66.0% of participants were females. During a mean follow-up of 11.94 years, we observed an elevated risk of CVD among patients with studied psychiatry disorders, compared with matched unexposed individuals (hazard ratios [HRs]=1.16, 95% confidence interval [CI]: 1.14-1.19), especially during the first six months of follow-up (HR=1.59 [1.42-1.79]). To assess the modification role of disease susceptibility, we stratified analyses by family history of CVD and by CVD PRS, which obtained similar estimates between subgroups with different susceptibilities to CVD. We conducted trajectory analysis to visualize the temporal pattern of CVDs after common psychiatry disorders, identifying primary hypertension, acute myocardial infarction, and stroke as three main intermediate steps leading to further increased risk of other CVDs.

**Conclusions:** The association between common psychiatry disorders and subsequent CVD is not modified by predisposition to CVD. Hypertension, acute myocardial infarction, and stroke are three initial CVDs linking psychiatric disorders to other CVD squeals, highlighting a need of timely intervention on these targets to prevent further CVD squeals among all individuals with common psychiatric disorders.

## Introduction

Both psychiatry disorders and cardiovascular disease (CVD) are leading causes of disability, morbidity, and mortality worldwide according to the World Health Organization (WHO)(DALYs et al., 2015; Whiteford et al., 2013). During the last decades, accumulating evidence indicate a link between psychiatry disorders and CVD(Han et al., 2021; Kollia et al., 2017; Momen et al., 2020; Song et al., 2019). Specifically, elevated risks of multiple CVDs, including ischemic heart disease(Janszky, Ahnve, Lundberg, & Hemmingsson, 2010; Wium-Andersen et al., 2019), hypertension(Stein et al., 2014), heart failure(Song et al., 2019), atherosclerosis(Yu, Ho, Lam, Woo, & Ho, 2010), and stroke(Momen et al., 2020; Swain et al., 2015), have been reported among patients with psychiatry disorders. Potential mechanisms linking psychiatric disorders and cardiovascular abnormalities are multifactorial, including chronic inflammation(Miller, Maletic, & Raison, 2009), alterations in hypothalamic– pituitary–adrenal (HPA) axis(Jokinen & Nordstrom, 2009), and unhealthy lifestyle (e.g., smoking)(Weinberger et al., 2017).

Recent genome wide association studies (GWAS) have suggested a genetic predisposition to different CVDs such as coronary artery disease(Consortium et al., 2013; Nikpay et al., 2015) and stroke(Network & International Stroke Genetics, 2016), including multiple single nucleotide polymorphisms (SNPs) significantly associated with these traits. Likewise, familial factors, as a proxy for the joint impact of genetic background and shared environmental and lifestyle factors, have been shown to play an important role in the development of CVD(Forsdahl, 1979; Iannotti, Zuckerman, & Rifai, 2000). However, data are scarce regarding whether predisposition to CVD can modify the association between psychiatry disorders and subsequent CVD. Findings from a Danish twin study did not support a role of shared familial factors on the associations of depression with CVD(Wium-Andersen et al., 2020). In our previous study, however, we noted attenuated risk increase of CVD among patients with stress-related disorders when comparing them with their full siblings, relative to comparison with age- and sex-matched unexposed individuals from the general populations(Song et al., 2019). These inconsistent results call for further investigations of this question, ideally with direct assessment on disease susceptibility to CVD such as individual-level genotype data.

Moreover, as there are shared etiologies and comorbidity between different CVDs(Collaboration, 2015; Laurent & Boutouyrie, 2015), a further question is whether there are specific CVDs that more directly related to psychiatric disorders and may lead to other CVDs downstream. We therefore applied disease trajectory analysis to map out the temporal pattern of CVDs after psychiatric disorders (Han et al., 2021; Siggaard et al., 2020). As the illustration of the progression network of CVDs can aid the identification of key CVDs first affected by psychiatric disorders, results of such analysis can be of importance in terms of helping identify targets for preventive measures with the goal of reducing the risk of subsequent CVDs.

In the present study, taking advantage of enriched phenotypic information, complete follow-up and individual-level genotyping data in the UK Biobank, we aimed to clarify whether disease susceptibility to CVD modifies the association of common psychiatry disorders with subsequent CVDs. Further, we used trajectory network analysis to identify the key CVDs directly associated with a prior diagnosis of common psychiatric disorders.

## Methods

### Study design

UK biobank is a community-based cohort study which enrolled a half million participants aged 40 to 69 years at recruitment between 2006 and 2010 across England, Scotland, and Wales. Baseline questionnaires and biological samples were collected at recruitment with detailed description elsewhere(Sudlow et al., 2015). Health-related outcomes were obtained by linkages to a range of health records(Sudlow et al., 2015). UK Biobank inpatient hospital data were mapped from the Hospital Episode Statistics database, the Scottish Morbidity Record, and the Patient Episode Database, covering 89% of UK Biobank participants since 1997. The primary care data were derived from various general practitioner computer system suppliers, providing information on primary care visit for ∼45% participants since 1985(Sudlow et al., 2015).

Based on these data, we conducted a matched cohort study. After the exclusion of 19 individuals who withdrew their informed consent forms, and 3 individuals who lost to follow-up before a diagnosis of common psychiatry disorders, we identified an exposed cohort of 39,960 participants who received their first diagnosis of anxiety, depression, or stress-related disorders between 1 January 1997 and 31 December 2019 (**Figure 1**). Individuals with those common psychiatry disorders were included in the exposed cohort from the date of their diagnosis (i.e., the index date). We then excluded individuals with a history of any CVD before the index date (n=4,733), leaving 35,227 exposed individuals in the final analysis. For each exposed individual (i.e., the index patient), we randomly selected five unexposed individuals per index patient, individually matched by sex and birth year (±2 years), from individuals who were free of those common psychiatry disorders and CVD at the diagnosis date of the index patient.

**Figure 1.**
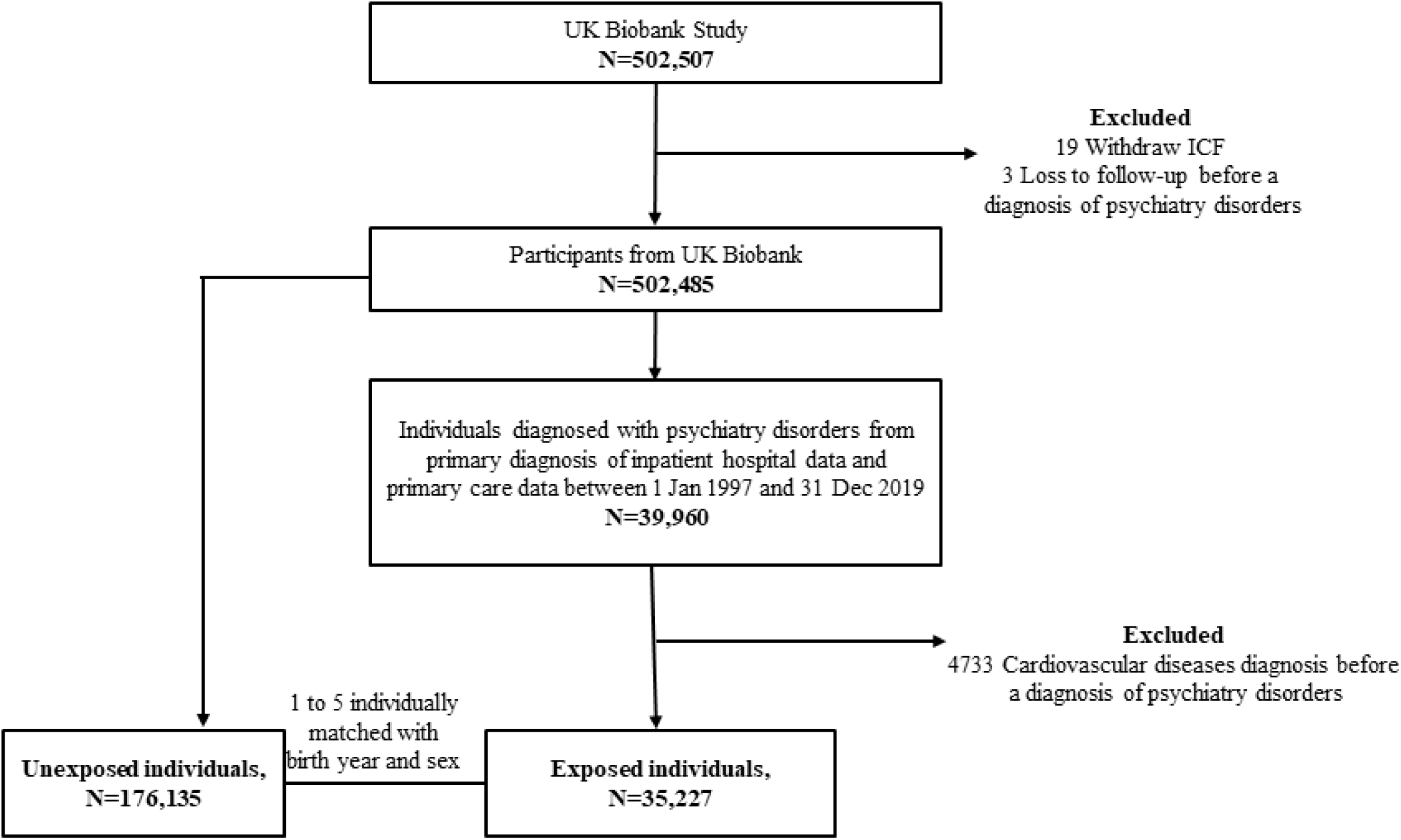
Flow chart the participants selection This figure shows the study design, the inclusion and exclusion process of study population selection.

#### Follow-up

For all participants, follow-up was from the index date until the diagnosis of any or specific CVDs, death, loss to follow-up, or the end of follow-up (31 December 2019), whichever occurred first. The follow-up for matched unexposed individuals was additionally censored at the time of their first diagnosis of studied psychiatry disorders, if any, during the follow-up.

#### Ascertainment of common psychiatric disorders

Common psychiatry disorders were defined as a diagnosis of anxiety, depression, or stress-related disorders, based on the primary diagnoses from UK Biobank inpatient hospital and primary care data. **eTable 1 in the supplement** lists details of diagnosis codes, including the 9^th^ to 10^th^ versions of the International Classification of Diseases (ICD) codes for inpatient hospital data, and version 2 and version 3 read codes (i.e., Read v2 and Read v3) for primary care data. We considered a diagnosis of other psychiatry disorders before the index date as history of other psychiatry disorders. Inpatient mental health diagnosis from the Hospital Episode Statistics database was validated with a median accuracy of 73% against a gold-standard research diagnosis(Davis et al., 2018), while the positive predictive value (PPV) of Read codes for anxiety and depression from primary care data was 61%-76% against the five-item Mental Health Inventory(John et al., 2016).

#### Ascertainment of CVD

With a specific focus on severe CVD consequences that require hospital care, we defined an incident CVD (any or six major subtypes including hypertensive diseases, ischemic heart diseases, embolism and thrombosis, arrhythmia/conduction disorders, heart failure, and cerebrovascular disease) as a corresponding diagnosis documented in the UK Biobank inpatient hospital data, according to ICD-9 and ICD-10 codes (**eTable 1 in the supplement**). For disease trajectory analysis, we used the 3-digit ICD-10 codes for CVD identification based on the UK Biobank inpatient hospital data and combined CVDs with clinical or biological similarities(Han et al., 2021; Yang et al., 2019) (**eTable 2 in the supplement**). The inpatient diagnosis of CVDs in UK Biobank has been validated, showing a satisfactory accuracy (i.e., PPV= 72% for coronary heart disease and >90% for stroke(Kivimaki et al., 2017; Woodfield et al., 2015)).

#### Disease susceptibility of CVD

In the present study, we assessed the disease susceptibility to CVD in two ways. First, we defined family history of CVD by a self-reported history of heart diseases and stroke among first-degree relatives (i.e., father, mother and siblings), obtained from baseline questionnaires (Field ID: 20107, 20110, 20111). Second, a polygenic risk score (PRS) for CVD was calculated for 376,019 individuals with eligible genotyping data (6,099,107 SNPs available after a standard quality control(Marees et al., 2018)) using LDPred2(Prive, Arbel, & Vilhjalmsson, 2020), a method for PRS computation based on combination of summary statistics and a matrix of correlations between genetic variants. The independent SNPs associated with CVD were derived from meta-analyzed GWAS summary data from CARDIoGRAMplusC4D Consortium(Nikpay et al., 2015). In a validation step, the generated CVD PRS was shown to be significantly associated with an increased risk of the CVD phenotype in our dataset (odds ratio [95% confidence intervals (CIs)]: 1.43 [1.40-1.45], per one unite increase of PRS).

#### Covariates

We retrieved information on educational levels, ethnicity, smoking status, Townsend deprivation index (TDI), and body mass index (BMI) through the baseline questionnaires. Based on the UK Biobank inpatient hospital data, we calculated the Charlson comorbidity index (CCI) on the index date for each individual, as a proxy of their baseline comorbidity level(Quan et al., 2005).

### Statistical analysis

We used Cox model to estimate hazard ratios (HRs) with 95% CIs of any CVD, in relation to a previous diagnosis of common psychiatry disorders, using time since the index date as the underlying time scale. The models were stratified by matched factors (sex and birth year), and partially or fully adjusted for educational levels, ethnicity, smoking status, TDI, BMI, history of other psychiatry disorders, family history of CVD, and CCI. As a high-risk time window for CVD has been indicated after diagnosis of psychiatric disorders(Song et al., 2019), we first visualized the changes of HRs over time by stratifying the Cox models to different follow-up periods (≤3, 3-6, 6-12, 12-18, 18-24, 24-60, 60-120, 120-240, and >240 months follow-up). As the HR during the first six months of follow-up period was greater than those of the later times (**eFigure 1 in the supplement**), we assessed the risk of CVD within and beyond the first six months (i.e., ≤6 or >6 months of follow-up) separately in later analyses. In addition, we did separate analyses for six main types of CVDs as well as for anxiety, depression and stress-related disorders.

To determine the impact of disease susceptibility to CVD on the association between common psychiatric disorders and CVD, we conducted stratified analyses by family history of CVD (yes or no) and by level of CVD PRS (high, moderate, or low by tertile distribution).

The temporal progression of CVD subsequent to a diagnosis of psychiatry disorders was explored using trajectory network analysis as described in our previous study(Han et al., 2021). In brief, a phenome-wide association analysis (PheWAS) was performed to identify specific CVDs associated with a prior diagnosis of psychiatry disorders. In this step, the p-value for statistical significance was set to 0.05/number of analyses performed (Bonferroni corrections), to account for multiple testing. We then used binomial tests to identify pairs of CVD events with a temporal order, and assessed the magnitude of the associations between CVD pairs through a nested case-control study design and conditional logistic regression (**eFigure 2 in the supplement**).

To test the robustness of our results to the definition of CVD, we reassessed these associations by identifying CVD solely based on the primary diagnosis from UK Biobank inpatient hospital data. All statistical analyses were conducted by R (version 4.0.2), PLINK (version 1.9), Python (version 3.8) and Cytoscape (version 3.8.0). Codes script used in the primary analyses are available (Source code 1).

## Results

In total, we identified 35,227 patients with studied common psychiatry disorders, together with 176,135 sex- and birth year-matched unexposed individuals (**Figure 1**). **Table 1** shows the basic characteristics of the study population. Mean age at index date was 51.76 years, and 66.0% of participants were females. Compared with matched unexposed individuals, patients with common psychiatry disorders were more likely to be smokers (either previous or current, 47.4% vs 41.6%), have a history of other psychiatric disorders (16.0% vs 4.1%), have lower educational level (41.0% vs 44.9% for university level), but higher BMI (26.6% vs 22.2% for more than 30 Kg/M^2^).

**Table 1.** Baselines characteristics of the study cohort

|  | **Total individuals** | **Individuals with**<br>**psychiatry**<br>**disorders**<br>**(n=35,227)** | **Matched**<br>**individuals without**<br>**psychiatry**<br>**disorders<sup>b</sup>**<br>**(n=176,135)** |
| --- | --- | --- | --- |
| **Follow-up time**<br>**(years), mean (SD)** | 11.94 (5.76) | 11.86 (5.81) | 11.95 (5.75) |
| **Age at index date,**<br>**mean (SD)** | 51.76 (8.96) | 51.72 (9.00) | 51.77 (8.96) |
| **Sex (%)** |  |  |  |
| Female | 139446 (66.0) | 23241 (66.0) | 116205 (66.0) |
| Male | 71916 (34.0) | 11986 (34.0) | 59930 (34.0) |
| **Educational levels**<br>**(%)** |  |  |  |
| University | 93568 (44.3) | 14452 (41.0) | 79116 (44.9) |
| High school | 24712 (11.7) | 3926 (11.1) | 20786 (11.8) |
| Middle school | 59307 (28.1) | 10219 (29.0) | 49088 (27.9) |
| Unknown | 33775 (16.0) | 6630 (18.8) | 27145 (15.4) |
| **Ethnicity (%)** |  |  |  |
| White | 189912 (89.9) | 32142 (91.2) | 157770 (89.6) |
| Others | 20304 (9.6) | 2899 (8.2) | 17405 (9.9) |
| Unknown | 1146 (0.5) | 186 (0.5) | 960 (0.5) |
| **BMI (%)** |  |  |  |
| < 18.5 | 1320 (0.6) | 216 (0.6) | 1104 (0.6) |
| 18.5-25 | 74888 (35.4) | 11180 (31.7) | 63708 (36.2) |
| 25-30 | 85499 (40.5) | 14257 (40.5) | 71242 (40.4) |
| > 30 | 48488 (22.9) | 9353 (26.6) | 39135 (22.2) |
| Unknown | 1167 (0.6) | 221 (0.6) | 946 (0.5) |
| **Smoking status**<br>**(%)** |  |  |  |
| Never | 120348 (56.9) | 18319 (52.0) | 102029 (57.9) |
| Previous smokers | 66836 (31.6) | 11593 (32.9) | 55243 (31.4) |
| Current smokers | 22991 (10.9) | 5101 (14.5) | 17890 (10.2) |
| Unknown | 1187 (0.6) | 214 (0.6) | 973 (0.6) |
| **Townsend**<br>**deprivation index<sup>a</sup>,**<br>**mean (SD)** | -1.25 (3.10) | -1.02 (3.16) | -1.29 (3.09) |
| **History of other**<br>**psychiatry**<br>**disorders (%)** |  |  |  |
| No | 198511 (93.9) | 29594 (84.0) | 168917 (95.9) |
| Yes | 12851 (6.1) | 5633 (16.0) | 7218 (4.1) |
| **Family history of** |  |  |  |
| **CVD (%)** |  |  |  |
| No | 93808 (44.4) | 14973 (42.5) | 78835 (44.8) |
| Yes | 117554 (55.6) | 20254 (57.5) | 97300 (55.2) |
| **CVD polygenic risk score** |  |  |  |
| Low | 54110 (25.60%) | 9243 (26.24%) | 44867 (25.47%) |
| Intermediate | 54110 (25.60%) | 9285 (26.36%) | 44825 (25.45%) |
| High | 54125 (25.61%) | 9400 (26.68%) | 44725 (25.39%) |
| Unknown | 49017 (23.19%) | 7299 (20.72%) | 41718 (23.69%) |
| **CCI score, mean (SD)** | 0.01 (0.19) | 0.01 (0.22) | 0.01 (0.19) |

This table shows the basic characteristics of the patients with common psychiatry disorders and their matched unexposed individuals.
Abbreviation: CCI: Charlson comorbidity index; SD: standard deviation; BMI: Body Mass Index; CVD: cardiovascular disease.
<sup>a</sup> Townsend deprivation index was assigned to each individual based on their postcode locations, with a greater index score implying a greater degree of deprivation; missing data of TDI (n=300) was replaced by mean TDI of all individuals.
<sup>b</sup> At most 5 participants who were alive, retained in the cohort and free of psychiatry disorders at the corresponding index date were individually matched to each individual with psychiatry disorders based on sex, and year of birth.

During a mean follow-up time of 11.94 years, 10,619 (crude incidence rate: 25.41 per 1000 person-years) and 43,192 (20.52 per 1000 person-years) incident cases of CVD were identified among the exposed and matched unexposed individuals, respectively. According to the fully adjusted Cox model, the risk of CVD was higher for patients with common psychiatry disorders compared with matched unexposed individuals (HR [95% CIs]: 1.16 [1.14-1.19]), especially during the first six months of follow-up (1.59 [1.42-1.79], **Table 2**). Except for heart failure, all other five main categories of CVD were associated with psychiatry disorders, with the strongest association observed for cerebrovascular disease (1.28 [1.19-1.38], **Table 2**). The result patterns were similar for the periods beyond six months of follow-up, while the highest HR was for heart failure within six months of follow-up (2.66 [1.42-4.97], **Table 2**). Comparable results were found for anxiety, depression, and stress-related disorders (**eTable 3 in the supplement**).

**Table 2.**
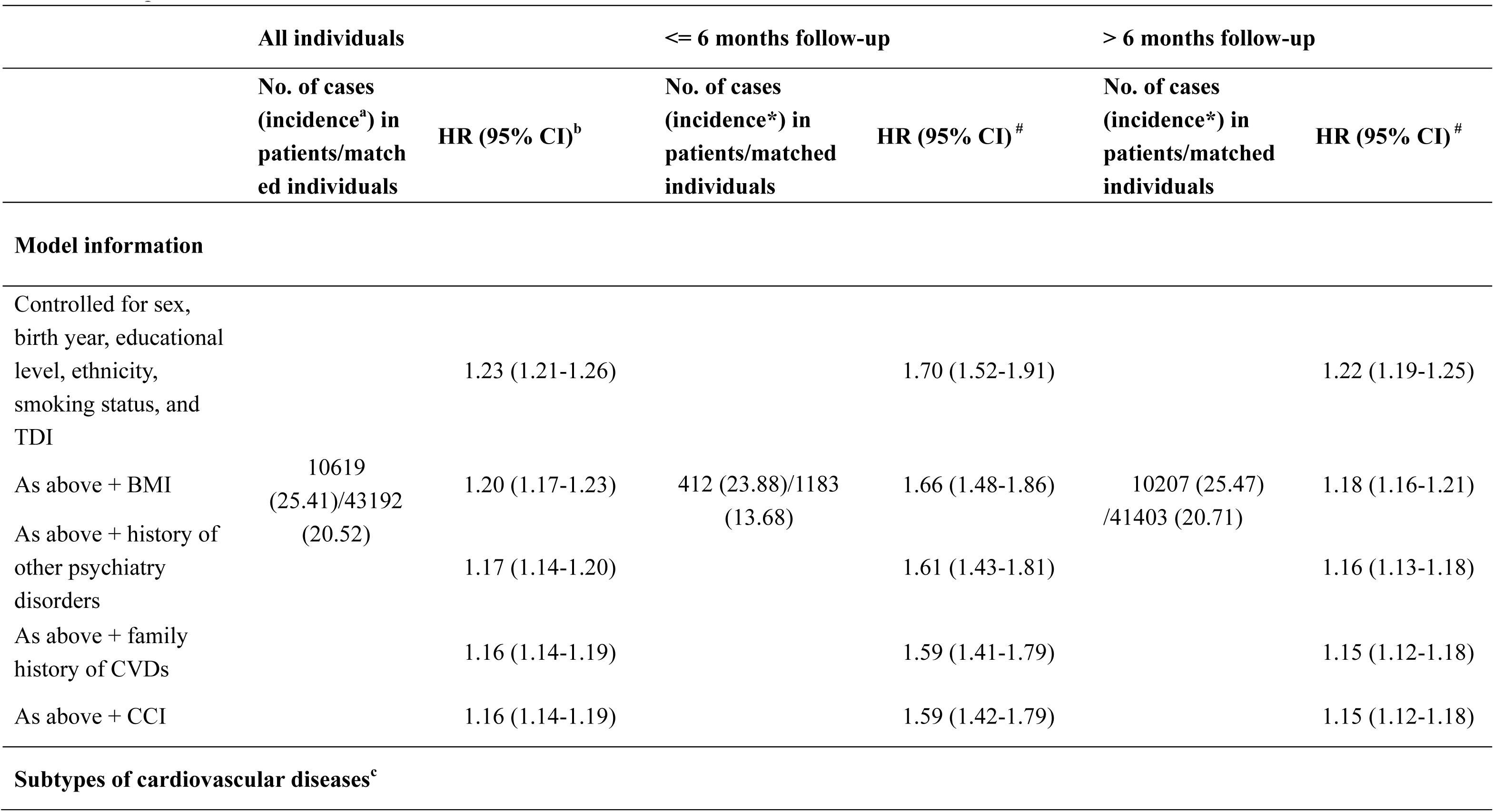
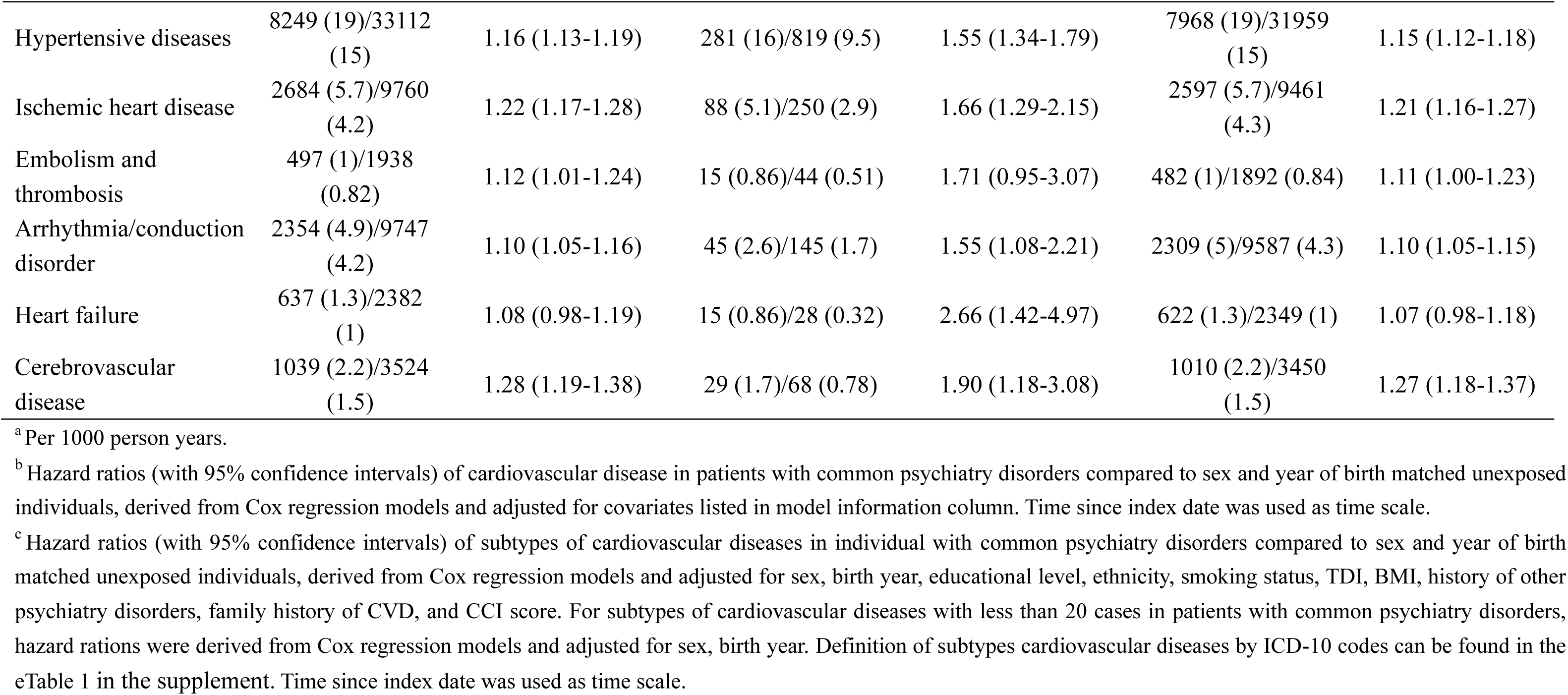
Hazard ratios (HRs) with 95% confidence intervals (CIs) of cardiovascular disease among patients with common psychiatry disorders compared to matched unexposed individuals

### The role of disease susceptibility

We observed similar associations among individuals with and without a family history of CVD (1.54 [1.31-1.80] vs 1.61 [1.25-2.07], P_interaction_ = 0.7703 within six months of follow-up, and 1.15 [1.11-1.18] vs 1.21 [1.16-1.27], P_interaction_ = 0.0683 beyond six months follow-up, **Table 3**). Likewise, comparable risk elevations of CVD were observed among individuals with different levels of CVD PRS, within six months of follow-up (2.29 [1.58-3.32] and 1.41 [0.98-2.02] for low and high PRS, P_interaction_ = 0.0667) and beyond (1.25 [1.16-1.34] and 1.18 [1.10-1.25] for low and high PRS, P_interaction_ = 0.2412) (**Table 3**). Analyses on subtypes of CVD by family history of CVD and CVD PRS showed similar result patterns (**eFigure 3 in the supplement)**.

**Table 3.**
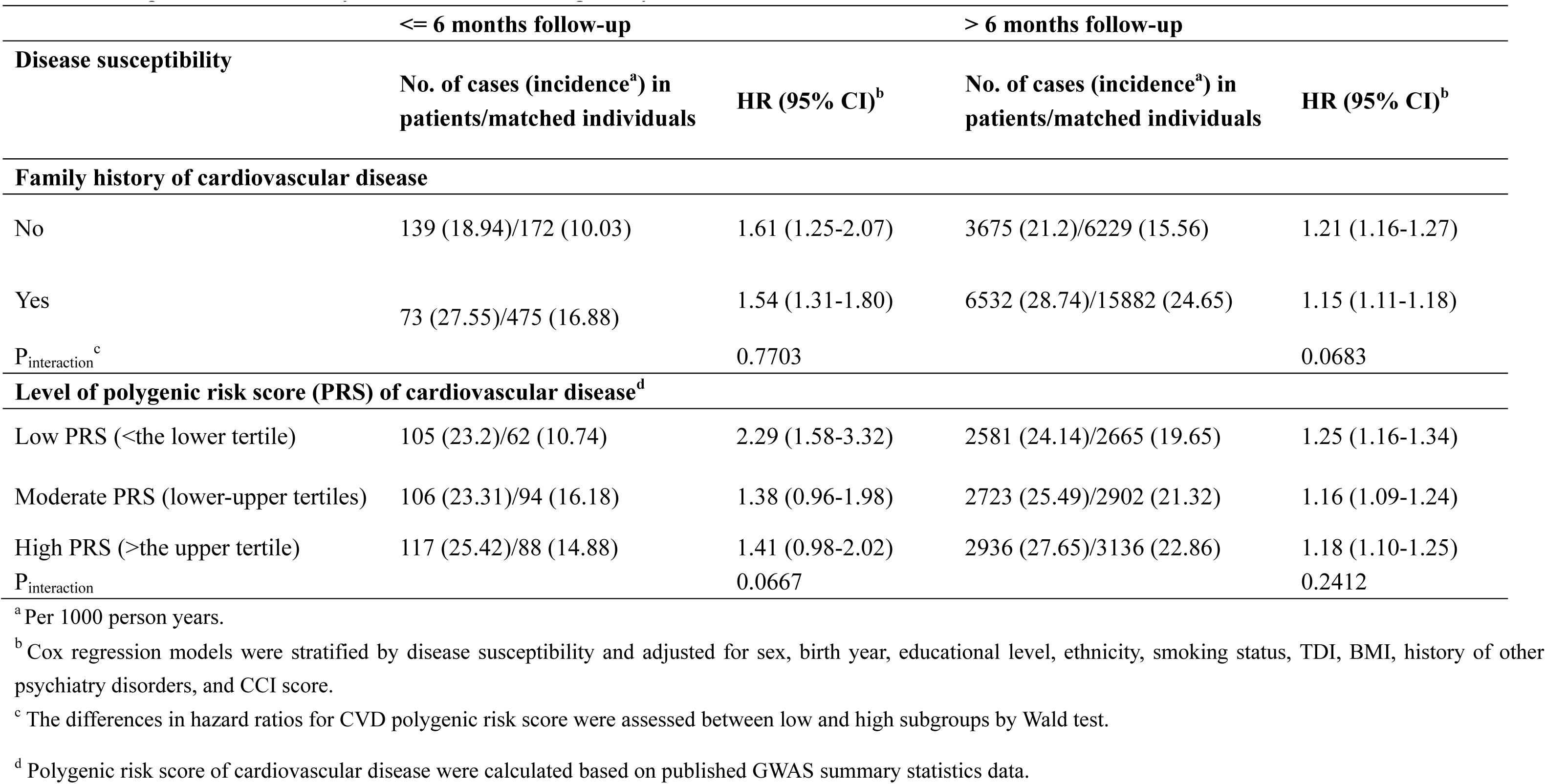
Hazard ratios (HRs) with 95% confidence intervals (CIs) of cardiovascular disease among patients with common psychiatry disorders compared to matched unexposed individuals, by different disease susceptibility

### Trajectory network analysis

15 CVDs with more than 150 cases among patients with common psychiatry disorders were involved in the PheWAS analysis (**eFigure 2 in the supplement**). Among these, eight CVDs showed significant associations with a prior diagnosis of common psychiatry disorders (**eTable 5 in the supplement**), forming 56 CVD pairs among which 21 passed the threshold of prevalence (i.e., experienced by at least 75 individuals). Finally, 12 pairs contributed to the visualized trajectory networks (**Figure 2** **and eTable 5 in the supplement**), presenting as a CVD disease tree originated from primary hypertension, acute myocardial infarction and stroke, which further connected to a wide range of other CVDs.

**Figure 2.**
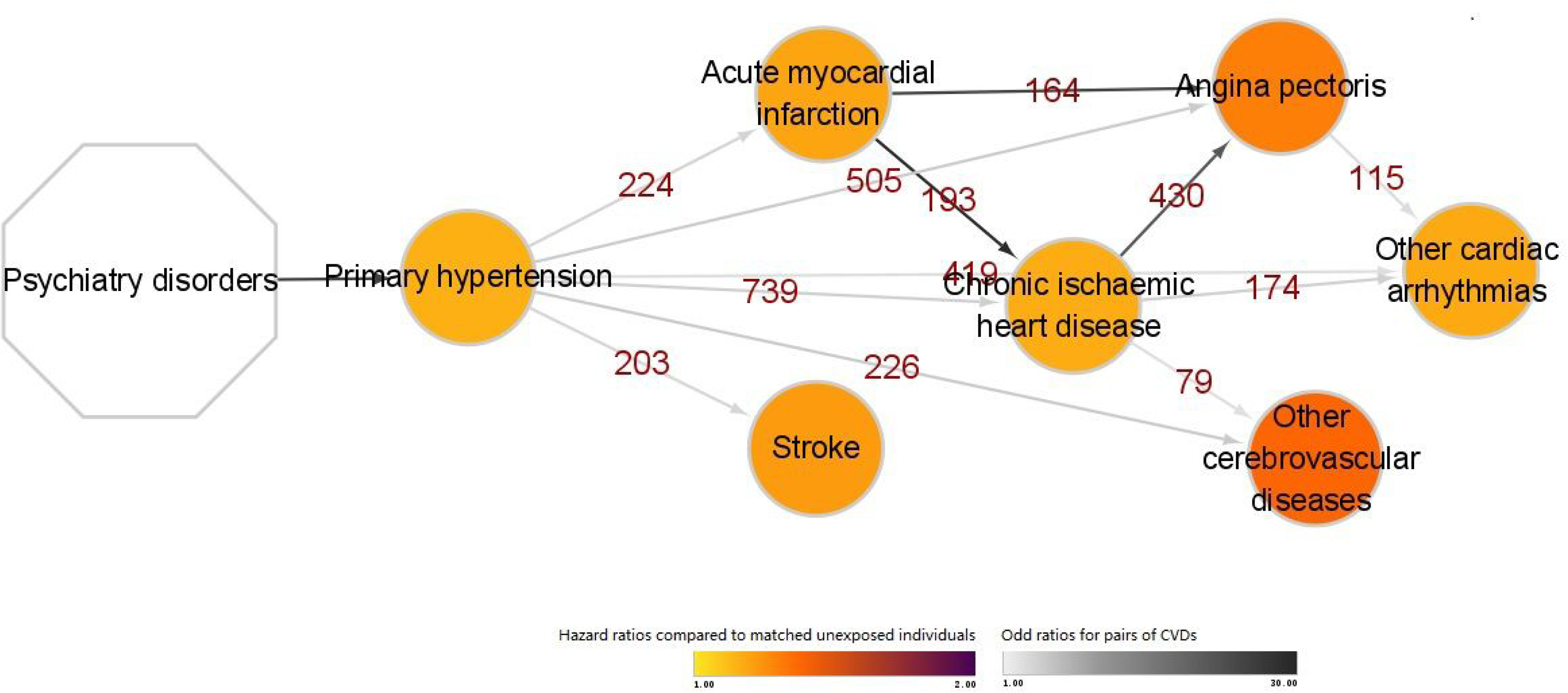
Trajectory progression of cardiovascular disease following a diagnosis of common psychiatry disorders This figure illustrates following trajectory progression of cardiovascular disease identified in our analysis. The combined cardiovascular diseases are shown within the circle. The color of the circle represents the hazard ratios of this cardiovascular disease when comparing patients with common psychiatry disorders to matched unexposed individuals. The number above the arrow connecting two circles corresponds to the number of pairs of two cardiovascular disease events among patients with common psychiatry disorders. The color of the arrows indicates the odds ratio of the sequential association between the two cardiovascular disease events among patients with common psychiatry disorders.

In the sensitivity analysis, defining the CVD cases solely by primary diagnoses, instead of all diagnoses, in UK Biobank inpatient data led to largely similar estimates (**Tables 6 and 7 in the supplement**). However, the number of CVDs involved in the trajectory network was substantially reduced, due to insufficient data power, where we had only 2 CVD pairs in the disease tree with an initial node of acute myocardial infarction (**eFigures 4 and 5 in the supplement**).

## Discussion

In this matched cohort study of UK Biobank, we found that individuals with common psychiatry disorders (i.e., anxiety, depression, and stress-related disorders) were at elevated risk of multiple CVDs, especially within first six months of follow-up. With mutually verified results obtained from stratified analyses by family history of CVD and by CVD PRS, the observed associations seemed to be constant across individuals with different predisposition to CVD. Furthermore, trajectory network analysis indicated that primary hypertension, acute myocardial infarction, and stroke were the CVD types firstly affected by a diagnosis of common psychiatric disorders which then further led to other CVDs downstream. This result highlights the need of timely intervention on these targets for preventing or interrupting further CVD squeals among individuals with common psychiatric disorders.

In line with our results, multiple large-scale cohort studies have reported an association between a diagnosis of psychiatry disorders and subsequently elevated risk of CVD, despite often insufficient control of important confounders, such as lifestyle(Han et al., 2021; Song et al., 2019; Stein et al., 2014; Wium-Andersen et al., 2019) and familial factors(Han et al., 2021; Kollia et al., 2017; Momen et al., 2020; Stein et al., 2014; Swain et al., 2015; Yu et al., 2010). Our finding that the association was not modified by family history of CVD gains support from two recent Danish twin studies(Wium-Andersen et al., 2020; Wium-Andersen et al., 2019), both of which concluded that genetic factors and early-life environment did not explain the association of depression with ischemic heart disease and stroke using a study sample of 6714 twins(Wium-Andersen et al., 2019) and 63,038 twins(Wium-Andersen et al., 2020), respectively. The Nurses’ Health Study II and the Normative Aging Study also revealed no attenuation in the magnitude of association between psychiatric disorders and CVD after additional adjustment for family history of CVD(Kubzansky, Koenen, Spiro, Vokonas, & Sparrow, 2007; Sumner et al., 2015). Our analysis on CVD PRS expanded this knowledgebase further and, for the first time, showed similar associations across groups with different levels of disease susceptibility to CVD. Taken together, these results underscore the importance of surveillance for CVD events among individuals with psychiatry disorders, regardless of disease liability.

Based on the trajectory network analysis, we identified three specific CVDs first affected by a prior diagnosis of common psychiatric disorders (i.e., primary hypertension, followed by acute myocardial infarction and stroke). Although there are no existing studies on CVD trajectory networks after psychiatry disorders, the direct effect of psychiatric disorders on specifically these CVDs has been implied in previous studies. Mendelian randomization analyses showed associations of genetic predisposition to depression with ischemic stroke(Cai et al., 2019) and myocardial infarction(Lu, Wang, Georgakis, Lin, & Zheng, 2021), suggesting a causal link between depression and these CVDs. Also, findings of experimental studies indicated direct physiological responses to mental stress, including endothelial dysfunction (monkeys(Strawn et al., 1991) and human(Ghiadoni et al., 2000)), dysregulation of blood pressure(Nas, Riese, van Roon, & Rijsdijk, 2020), and activated HPA axis(Jokinen & Nordstrom, 2009). Moreover, similar downstream CVDs (e.g., chronic ischemic heart diseases, other cerebrovascular diseases, and other cardiac arrythmias) after primary hypertension, and acute myocardial infarction can be identified using a disease trajectory browser that was developed based on national data from inpatient wards, outpatient clinics, and emergency room visits of Denmark(Siggaard et al., 2020). Collectively, these data suggest that these key CVDs may be potential targets for preventing or interrupting further CVD sequels among individuals with psychiatric disorders.

The major strength of our study includes the multidimensional data from UK Biobank, i.e., rich phenotypic information on family history of CVD, health-related outcomes, and individual-level genotype information, which enabled a comprehensive assessment on the modification role of CVD susceptibility on the studied associations and the temporal patterns of CVD development following a diagnosis of common psychiatry disorders. This rich data source also enabled vigorous control of a wide range of important confounders, including socio-demographic and behavioral factors, as well as baseline health status. Other strengths include the long and complete follow-up (mean follow-up time of 11.94 years), providing sufficient surveillance period for CVD outcomes, and the independent collections of diagnoses for psychiatry disorders and CVD, which minimizes the risk of information bias.

Our study has several limitations. First, as both primary and secondary diagnoses from UK Biobank inpatient hospital data were used for CVD identification, the diagnosis date for some mild forms of CVDs (e.g., primary hypertension) may not be accurate, leading to a risk of reverse causality. Consequently, the appearance of primary hypertension as the initial node in the CVD trajectory tree of the main analysis may rather imply the importance of primary hypertension as a comorbidity of psychiatric disorders on other subsequent CVDs, instead of a key progression pathway. Nevertheless, our analyses on acute and severe CVDs (e.g., acute myocardial infarction) should be less affected by such concern. Second, the SNPs used in the calculation for CVD PRS was obtained from GWAS studies which mainly focused on coronary arterial diseases, not all CVDs. However, a common genetic basis has been demonstrated between different CVDs(Dichgans et al., 2014; Jafaripour, Sasanejad, Dadgarmoghaddam, & Sadr-Nabavi, 2019), and the generated PRS has been confirmed to be associated with the CVD phenotype of our dataset in our validation step. Furthermore, we obtained largely similar result patterns in the analyses of six major categories of CVDs. Third, as the trajectory network analysis is a data-driven approach which requires a large sample size, our analysis might have missed some rare CVDs in the trajectory networks after psychiatric disorders. Further, because some important confounders, such as BMI and smoking status, were measured at baseline, they might not accurately reflect the real status of these factors at the time of psychiatric disorder diagnosis. Last, generalization of our findings to the entire UK population, or other populations, should be done with caution because the UK Biobank participants were overall healthier than the general UK polulation(Sudlow et al., 2015).

In conclusion, individuals with common psychiatry disorders have an elevated risk of CVD regardless of their predisposition to CVD. Hypertension, acute myocardial infarction, and stroke represent the three initial CVDs that link psychiatric disorders to further CVDs, highlighting a need of a timely intervention to prevent further CVD sequels among individuals with common psychiatric disorders.

## Data Availability

Source data for Figure 2 has been provided in the supplementary (eTable 5). Original Data is held by UK Biobank Limited. Due to the ethical approval of UK Biobank from the NHS National Research Ethics Service and the ethical approval of the current study, we cannot make the data publicly available. However, all researchers can access the data upon making an application to the UK Biobank. Detailed information on data application can be found at http://www.ukbiobank.ac.uk/.

## Acknowledgments

This research has been conducted using the UK Biobank Resource under Application 54803. We thank the team members and colleagues involved in West China Biomedical Big Data Center-UK Biobank project for their support.

## Contributions

HS, FF, and UAV were responsible for the study concept and design. XH, YH, YajingS, ZY, YQ, and JW did the data and project management. XH, YZ, HY, WC, and CH did the data cleaning and analysis. XH, YZ, HS, and FF interpreted the data. XH, YananS, UAV, FF, WZ, and HS drafted the manuscript. All authors approved the final manuscript as submitted and agree to be accountable for all aspects of the work. The corresponding author attests that all listed authors meet authorship criteria and that no others meeting the criteria have been omitted.

## Funding

This work is supported by the National Natural Science Foundation of China (No. 81971262 to HS), 1.3.5 project for disciplines of excellence, West China Hospital, Sichuan University (No. ZYYC21005 to HS), EU Horizon2020 Research and Innovation Action Grant (847776 to UV and FF).

## Disclosure

We declare no competing interests.

## Ethics approval

The UK Biobank has full ethical approval from the NHS National Research Ethics Service (reference number: 16/NW/0274), and this study was also approved by the biomedical research ethics committee of West China Hospital (reference number: 2019-1171).

## Additional information

Supplementary Tables and Figures are available at online website.

---

**eFigure 1.**
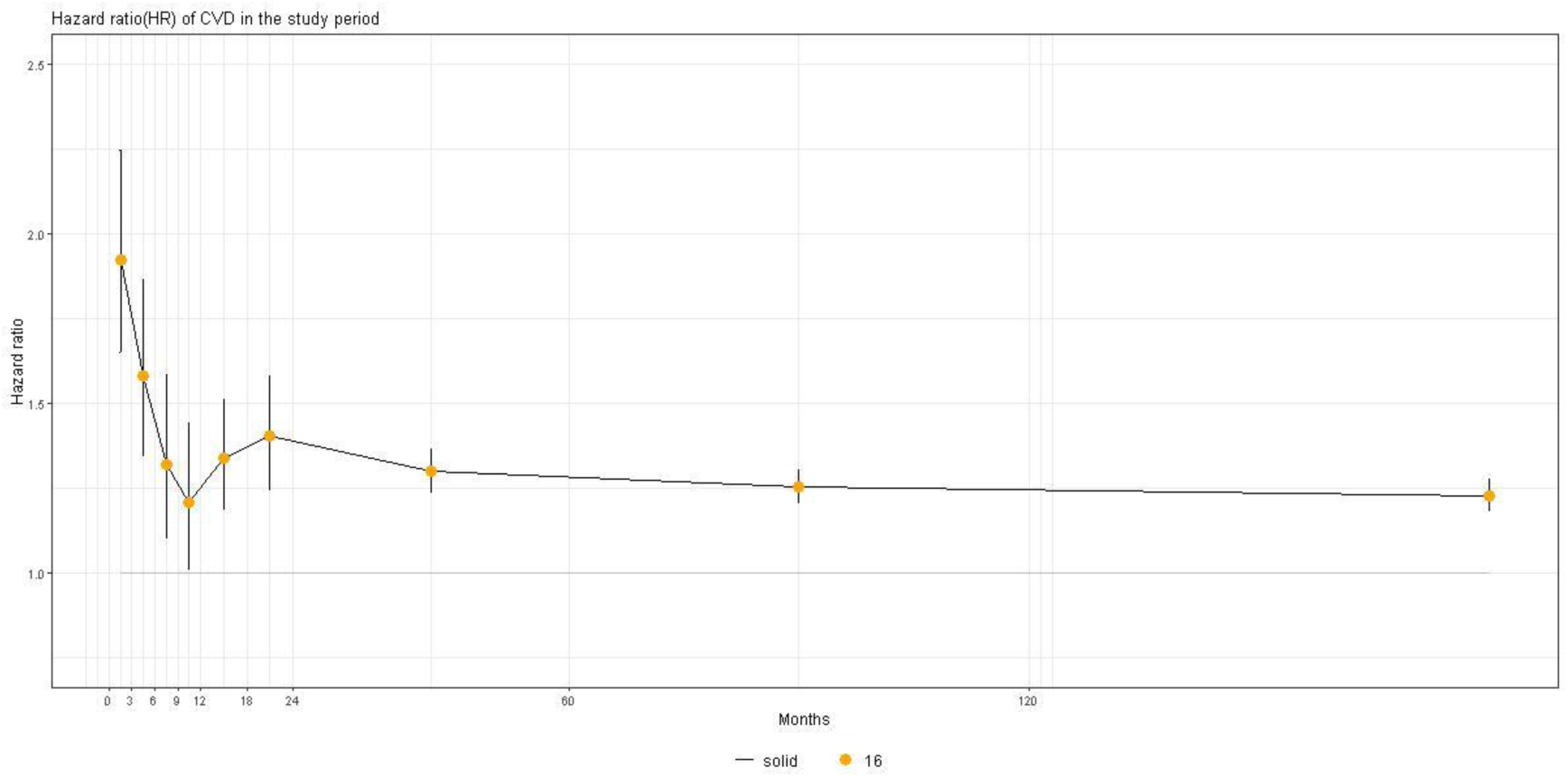
Hazard ratios (95% CIs) of cardiovascular disease among patients with common psychiatry disorders compared with their matched unexposed individuals, stratified by time of follow-up Abbreviation: CI: confidence interval. The X axis shows the different follow-up period: ≤3, 3-6, 6-12, 12-18, 18-24, 24-60, 60-120, 120-240, and >240 months follow-up. The Y axis shows the significant hazard ratios of cardiovascular disease among patients with common psychiatry disorders compared with their matched unexposed individuals, derived from Cox models adjusted for sex, birth year and ethnicity.

**eFigure 2.**
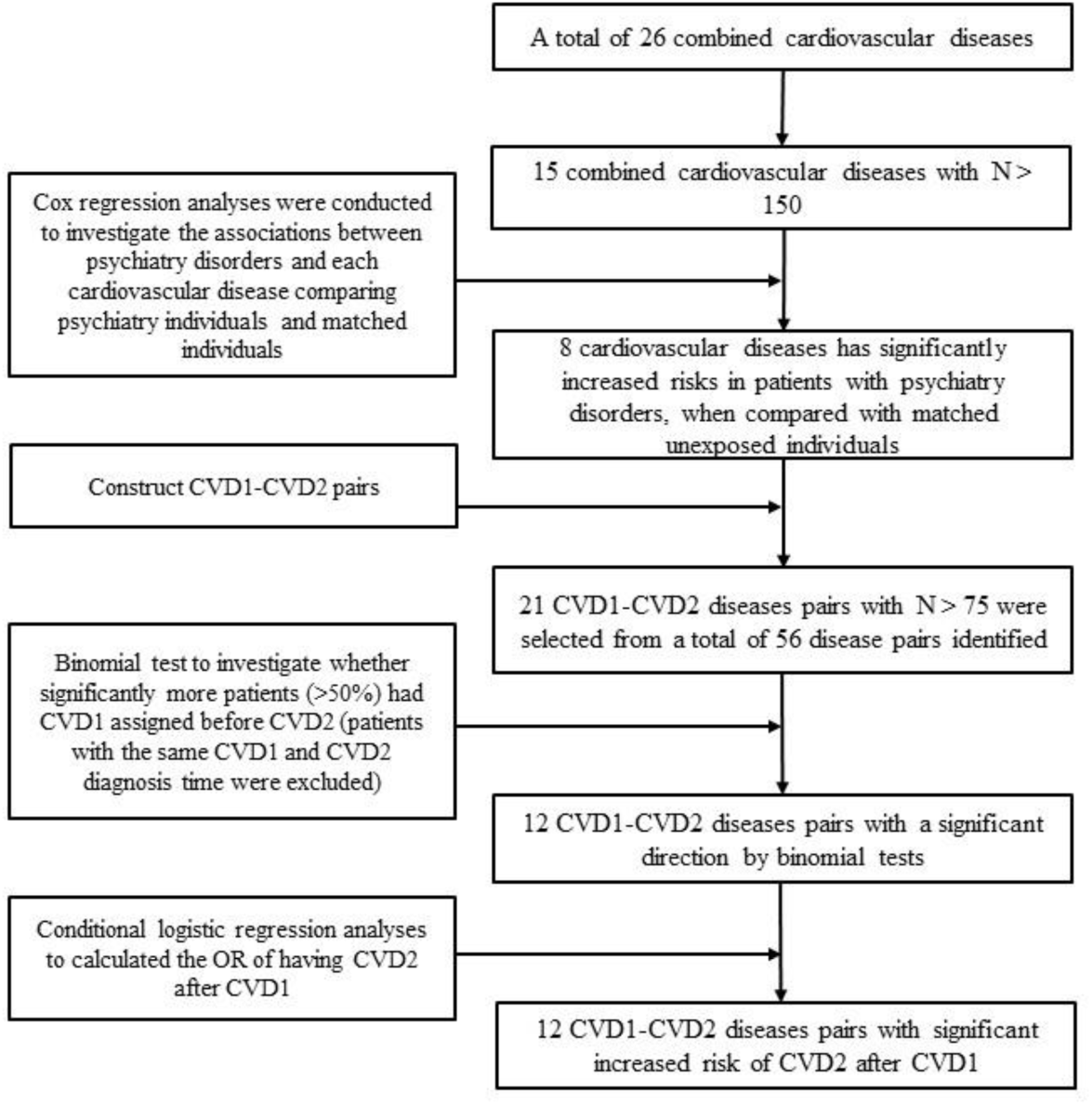
Flow chart of identifying trajectory progression of cardiovascular disease following a diagnosis of common psychiatry disorders

**eFigure 3.**
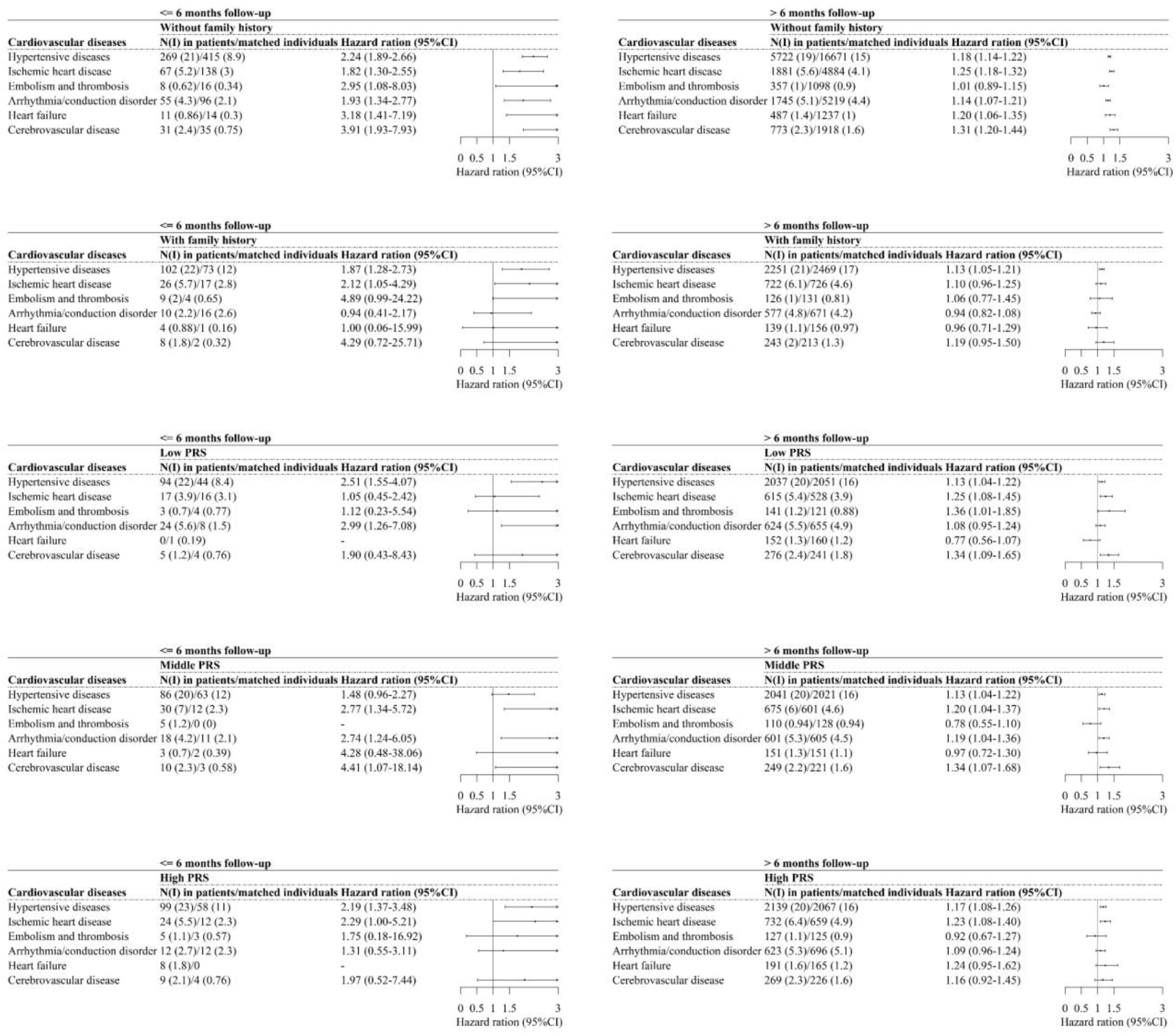
Relative risks of six subtypes of cardiovascular diseases among patients with common psychiatry disorders compared with their matched unexposed individuals, stratified by disease susceptibility, by time of follow-up (<= 6 or > 6 months) Abbreviation: N: number of cases of specific cardiovascular disease; I: incidence. Cox models were stratified by family history of cardiovascular disease (yes or no) and level of cardiovascular disease PRS (low, middle, and high) and adjusted for sex, birth year, educational level, ethnicity, smoke, TDI, BMI, history of other psychiatry disorders, and CCI score. For subtypes of cardiovascular diseases with less than 20 cases in patients with common psychiatry disorders, hazard rations were derived from Cox regression models and adjusted for sex, birth year.

**eFigure 4.**
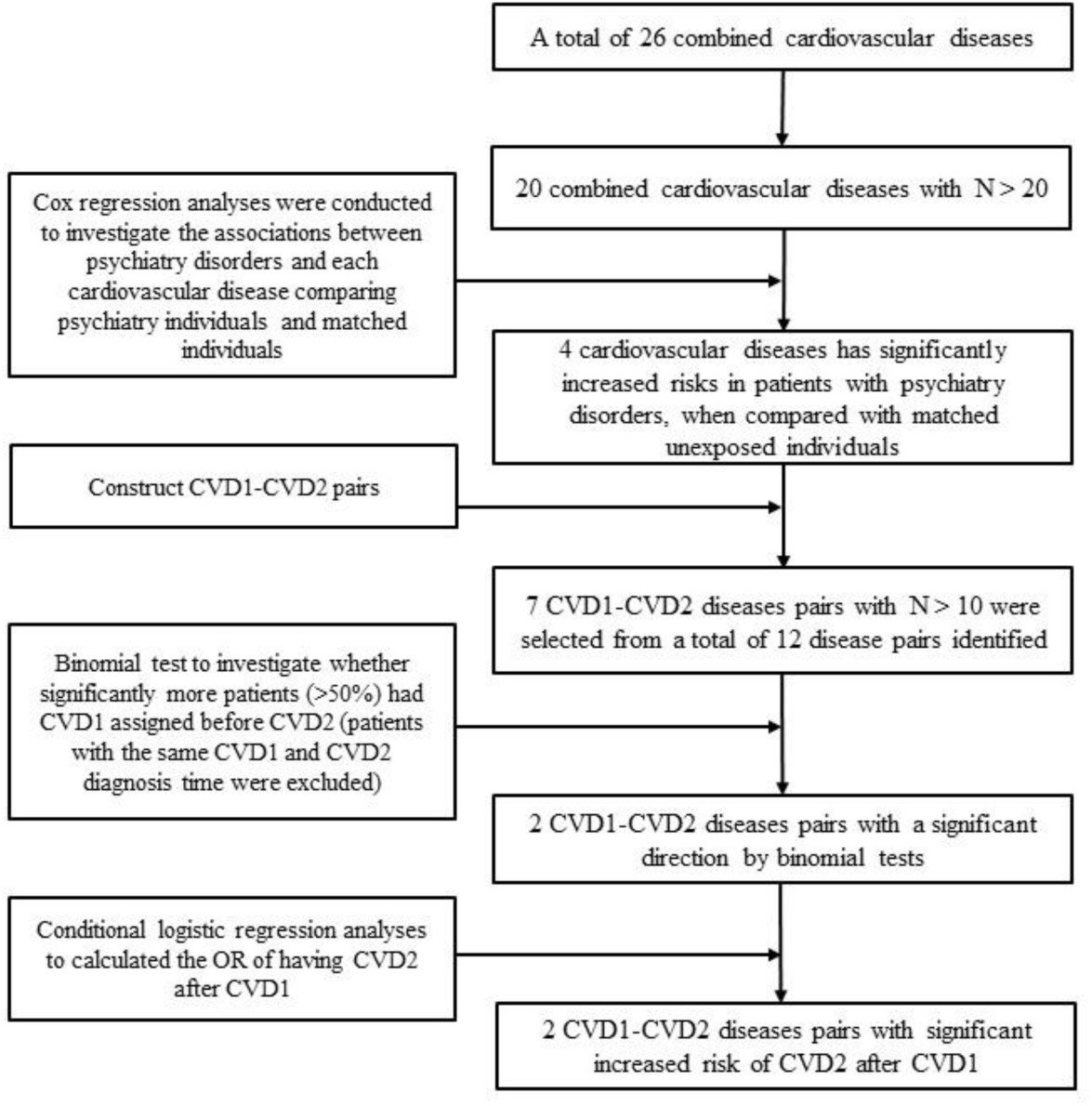
Flow chart of identifying trajectory progression of primary diagnosis of cardiovascular disease following a diagnosis of common psychiatry disorders

**eFigure 5.**
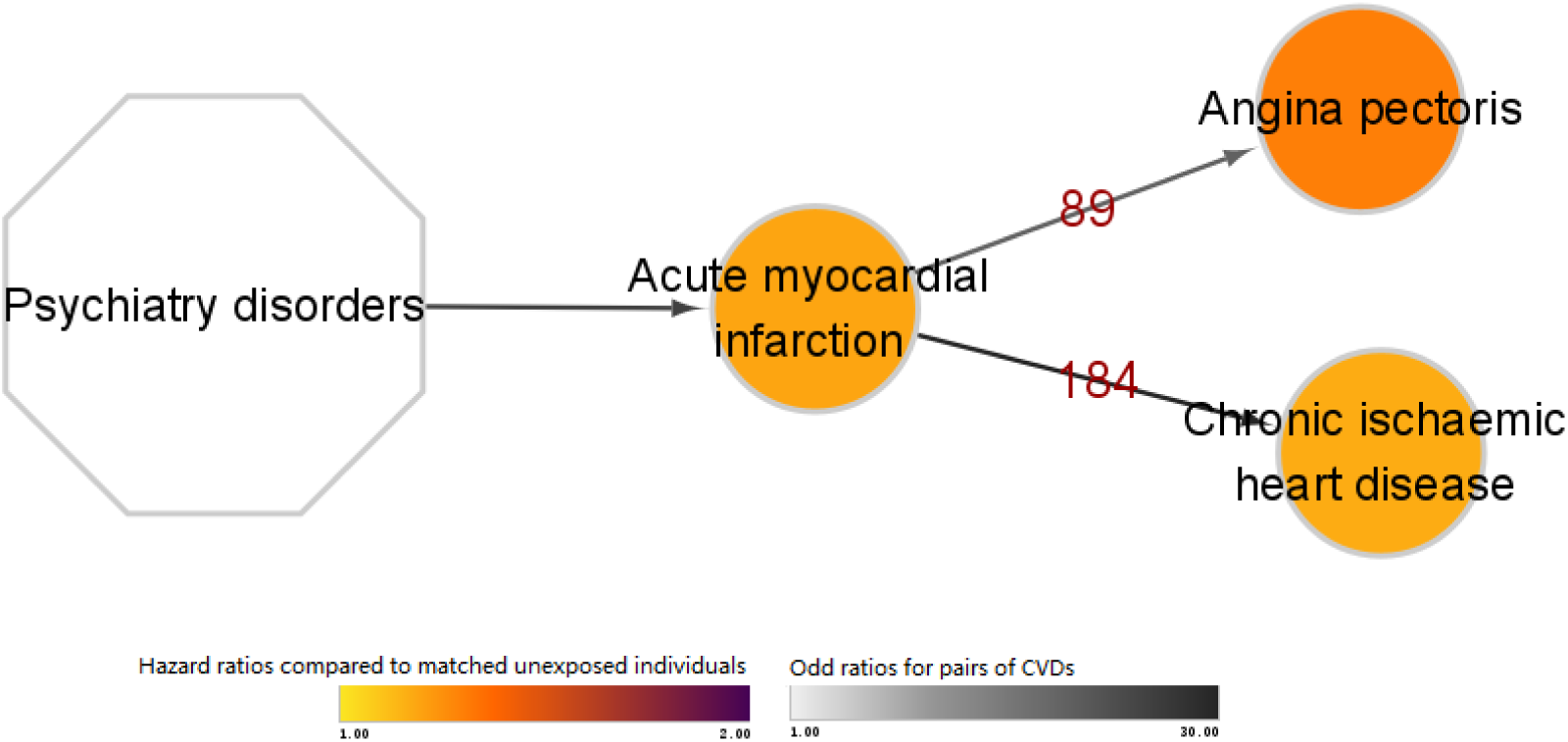
Trajectory progression of primary diagnosis of cardiovascular disease following a diagnosis of common psychiatry disorders This figure illustrates following trajectory progression of primary diagnosis of cardiovascular disease identified in our analysis. The combined cardiovascular diseases are shown within the circle. The color of the circle represents the hazard ratios of this cardiovascular disease when comparing patients with common psychiatry disorders to matched unexposed individuals. The number above the arrow connecting two circles corresponds to the number of pairs of two cardiovascular disease events among patients with common psychiatry disorders. The color of the arrows indicates the odds ratio of the sequential association between the two cardiovascular disease events among patients with common psychiatry disorders.

**eTable 1.**
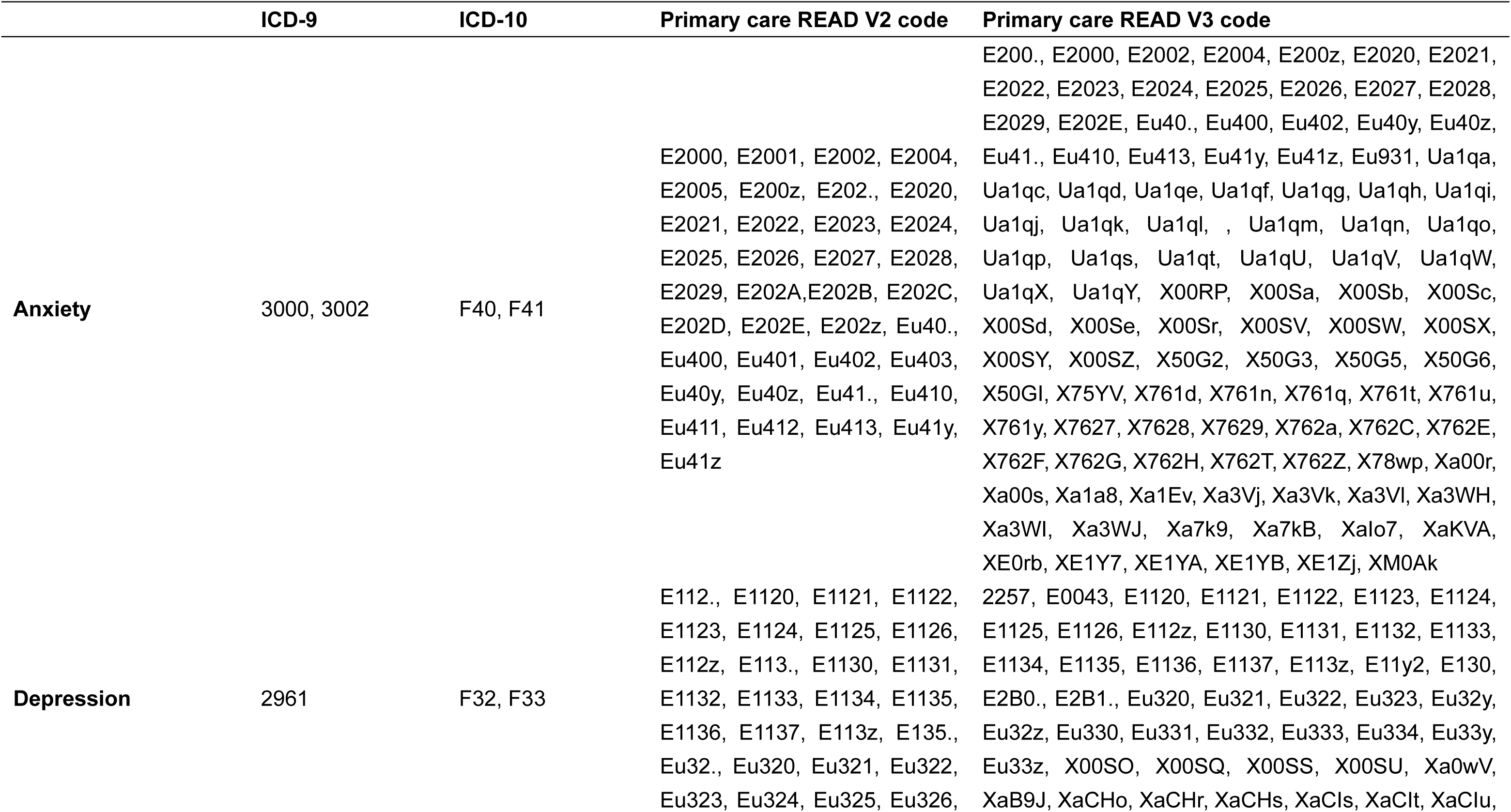
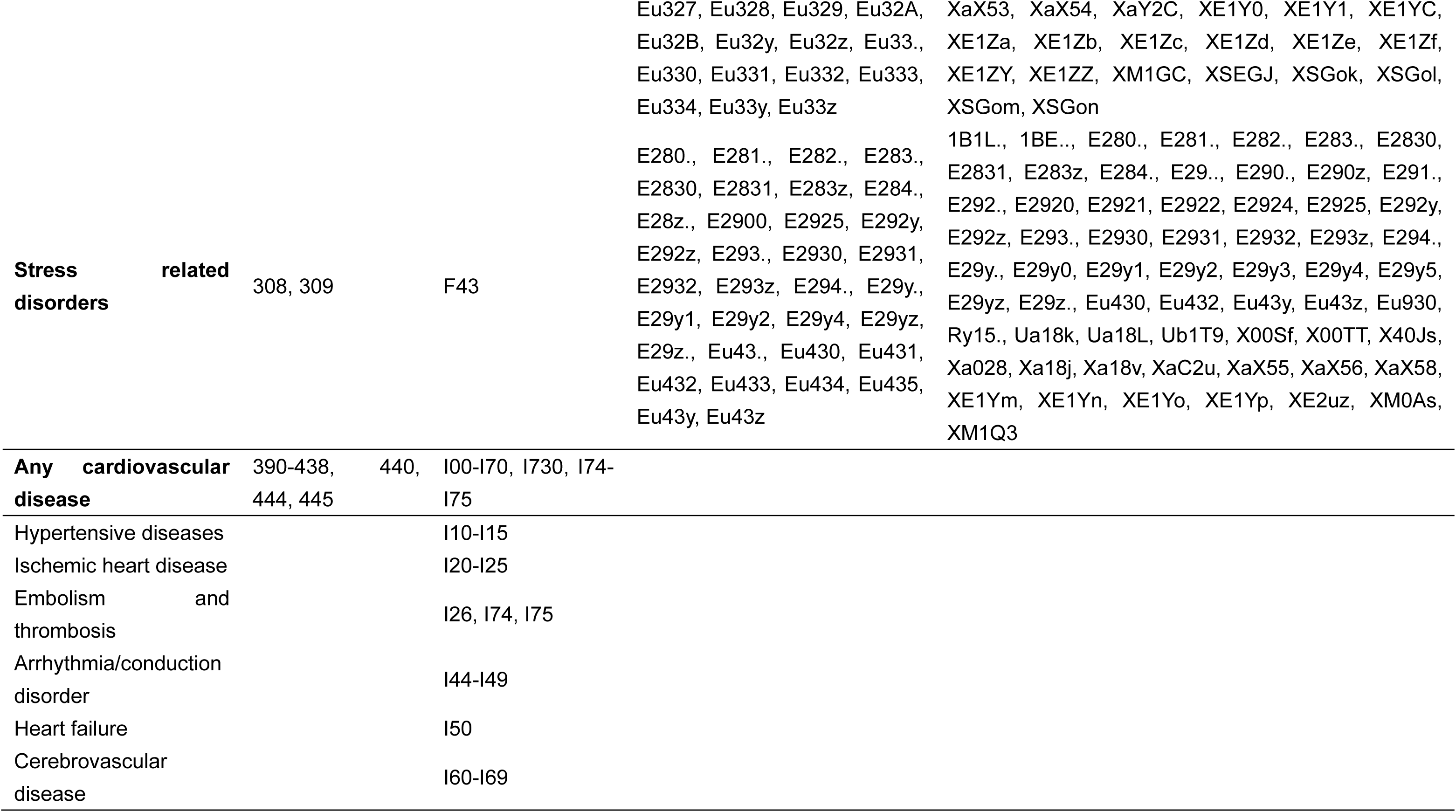
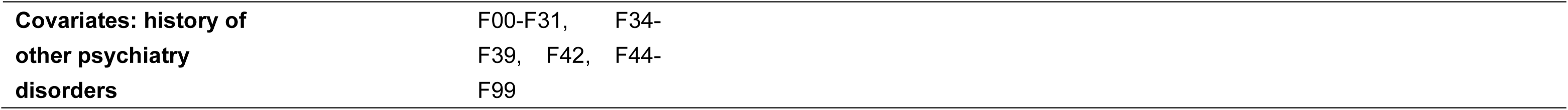
International Classification of Diseases (ICD) codes and primary care codes for diseases identifications

**eTable 2.**
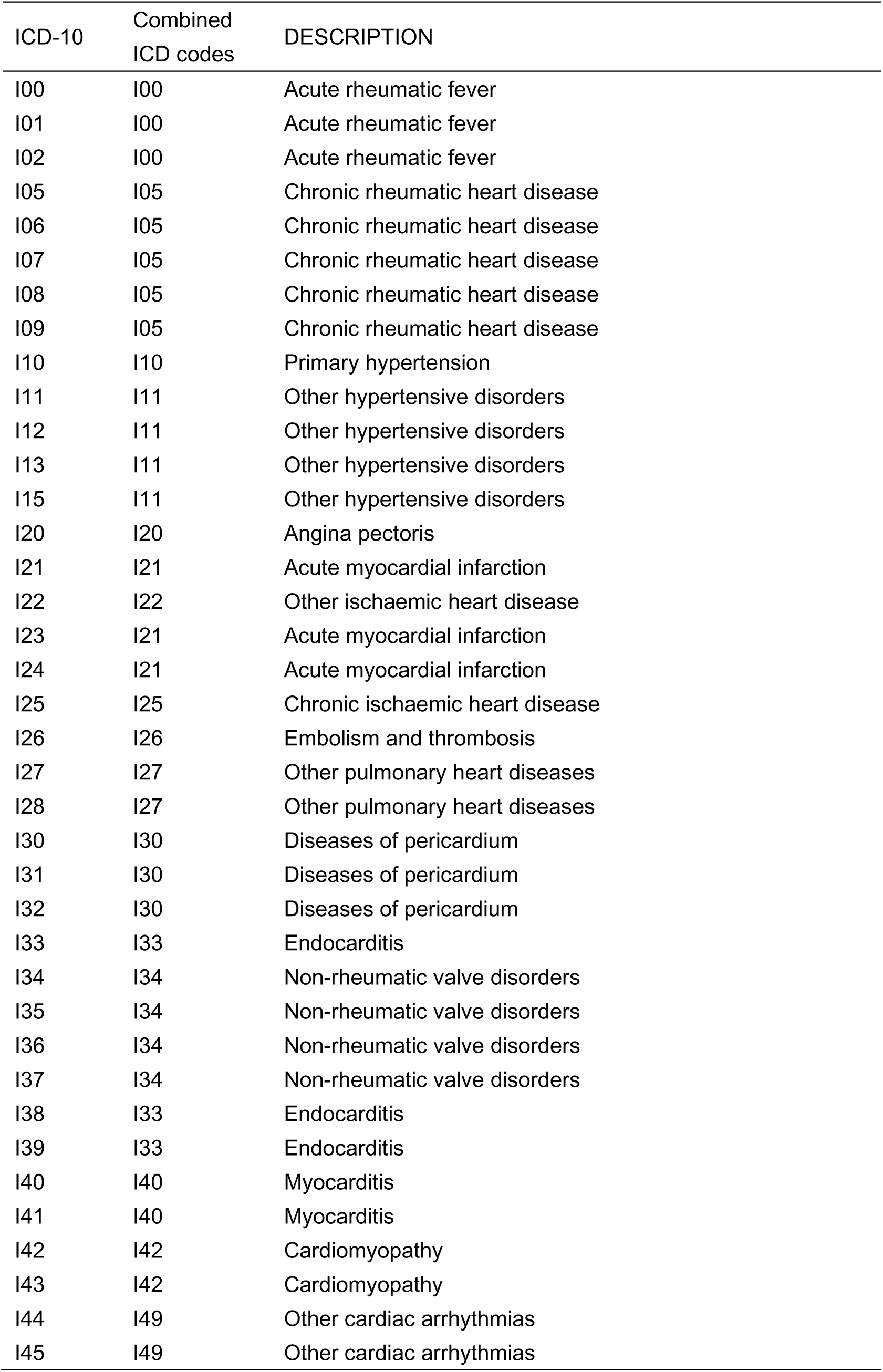
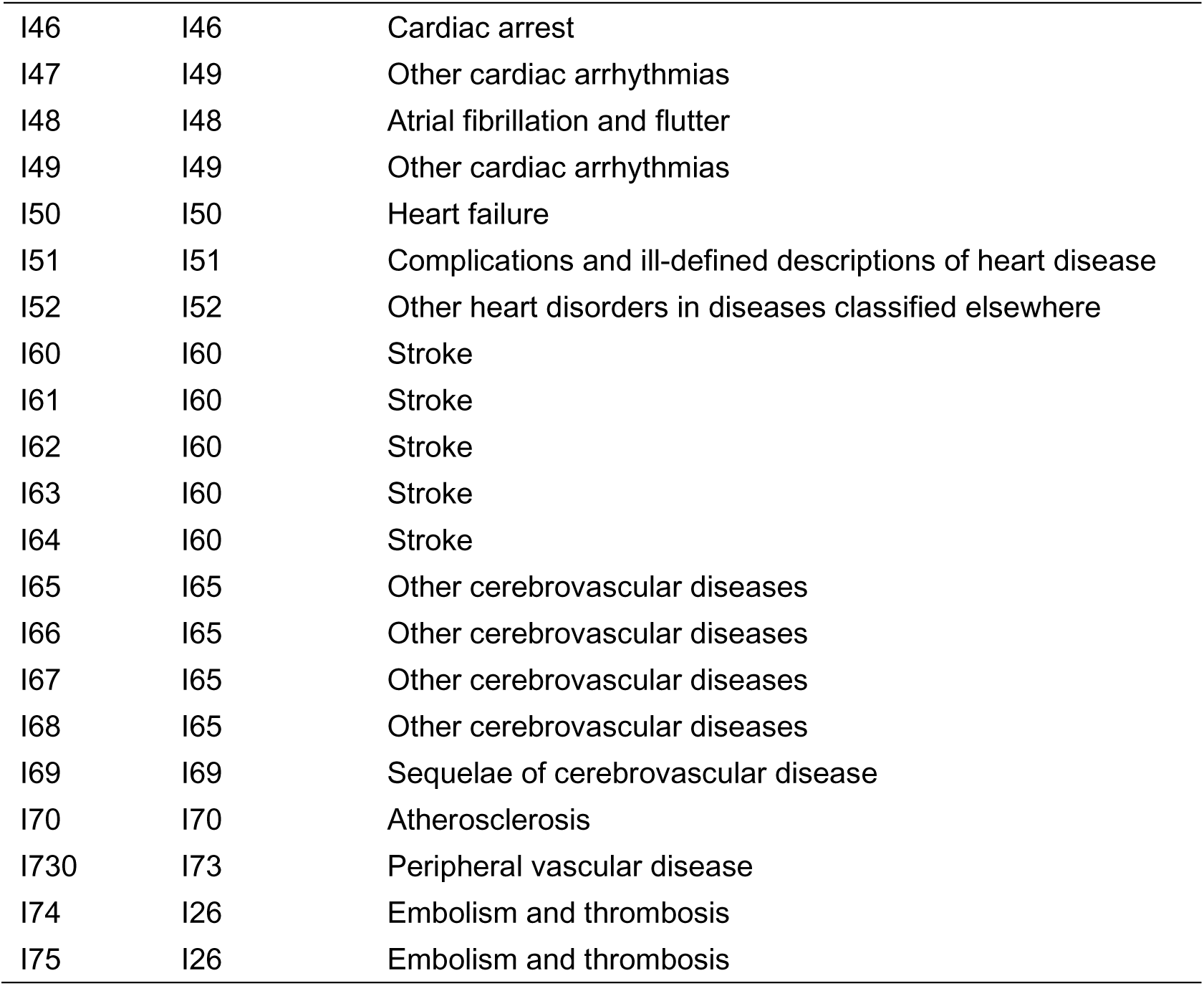
Mapping between ICD-10 codes and Combined ICD codes for PheWAS analysis

| ICD-10 | Combined ICD codes | DESCRIPTION |
| --- | --- | --- |
| I00 | I00 | Acute rheumatic fever |
| I01 | I00 | Acute rheumatic fever |
| I02 | I00 | Acute rheumatic fever |
| I05 | I05 | Chronic rheumatic heart disease |
| I06 | I05 | Chronic rheumatic heart disease |
| I07 | I05 | Chronic rheumatic heart disease |
| I08 | I05 | Chronic rheumatic heart disease |
| I09 | I05 | Chronic rheumatic heart disease |
| I10 | I10 | Primary hypertension |
| I11 | I11 | Other hypertensive disorders |
| I12 | I11 | Other hypertensive disorders |
| I13 | I11 | Other hypertensive disorders |
| I15 | I11 | Other hypertensive disorders |
| I20 | I20 | Angina pectoris |
| I21 | I21 | Acute myocardial infarction |
| I22 | I22 | Other ischaemic heart disease |
| I23 | I21 | Acute myocardial infarction |
| I24 | I21 | Acute myocardial infarction |
| I25 | I25 | Chronic ischaemic heart disease |
| I26 | I26 | Embolism and thrombosis |
| I27 | I27 | Other pulmonary heart diseases |
| I28 | I27 | Other pulmonary heart diseases |
| I30 | I30 | Diseases of pericardium |
| I31 | I30 | Diseases of pericardium |
| I32 | I30 | Diseases of pericardium |
| I33 | I33 | Endocarditis |
| I34 | I34 | Non-rheumatic valve disorders |
| I35 | I34 | Non-rheumatic valve disorders |
| I36 | I34 | Non-rheumatic valve disorders |
| I37 | I34 | Non-rheumatic valve disorders |
| I38 | I33 | Endocarditis |
| I39 | I33 | Endocarditis |
| I40 | I40 | Myocarditis |
| I41 | I40 | Myocarditis |
| I42 | I42 | Cardiomyopathy |
| I43 | I42 | Cardiomyopathy |
| I44 | I49 | Other cardiac arrhythmias |
| I45 | I49 | Other cardiac arrhythmias |
| I46 | I46 | Cardiac arrest |
| I47 | I49 | Other cardiac arrhythmias |
| I48 | I48 | Atrial fibrillation and flutter |
| I49 | I49 | Other cardiac arrhythmias |
| I50 | I50 | Heart failure |
| I51 | I51 | Complications and ill-defined descriptions of heart disease |
| I52 | I52 | Other heart disorders in diseases classified elsewhere |
| I60 | I60 | Stroke |
| I61 | I60 | Stroke |
| I62 | I60 | Stroke |
| I63 | I60 | Stroke |
| I64 | I60 | Stroke |
| I65 | I65 | Other cerebrovascular diseases |
| I66 | I65 | Other cerebrovascular diseases |
| I67 | I65 | Other cerebrovascular diseases |
| I68 | I65 | Other cerebrovascular diseases |
| I69 | I69 | Sequelae of cerebrovascular disease |
| I70 | I70 | Atherosclerosis |
| I730 | I73 | Peripheral vascular disease |
| I74 | I26 | Embolism and thrombosis |
| I75 | I26 | Embolism and thrombosis |

**eTable 3.**
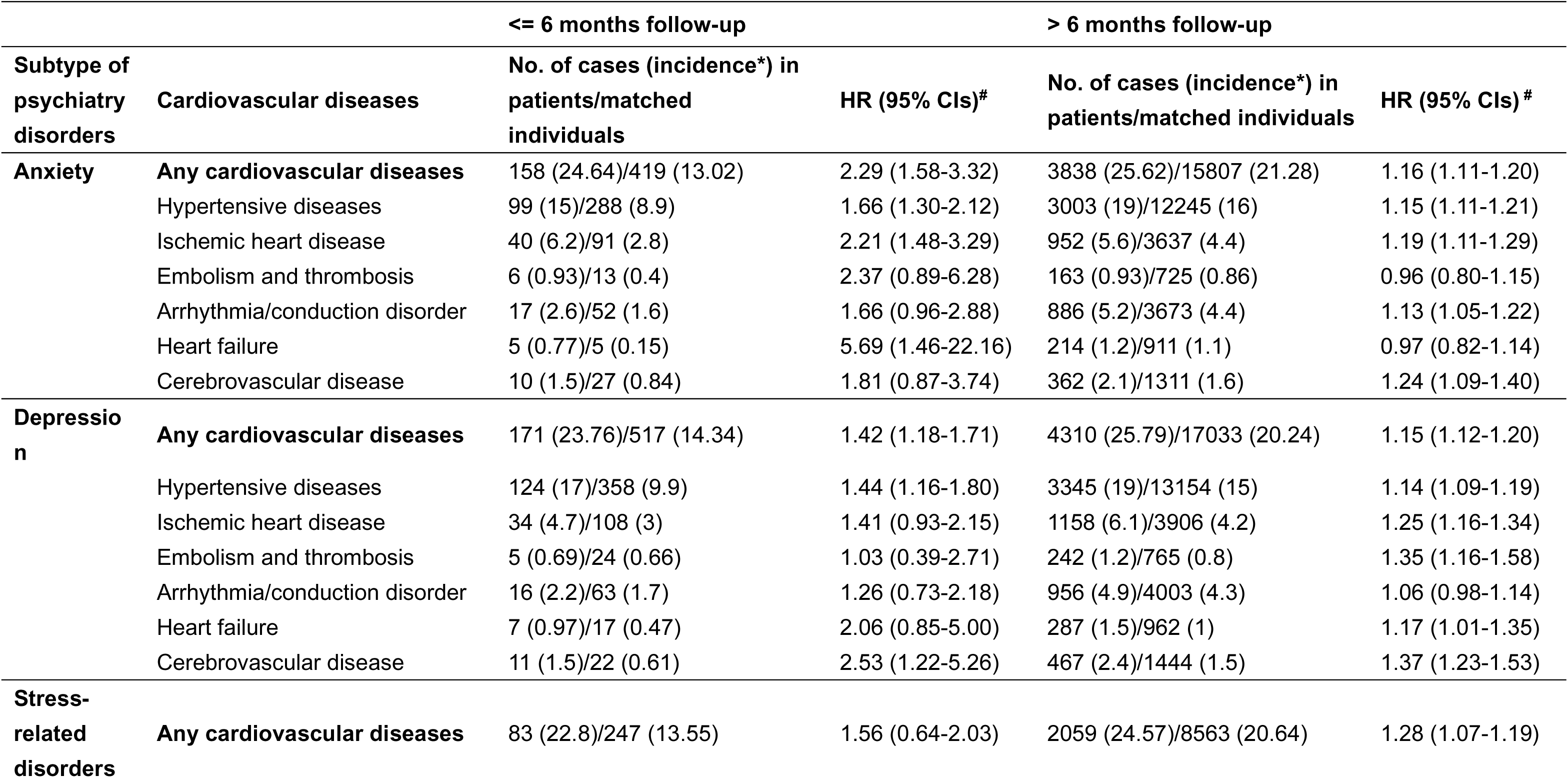
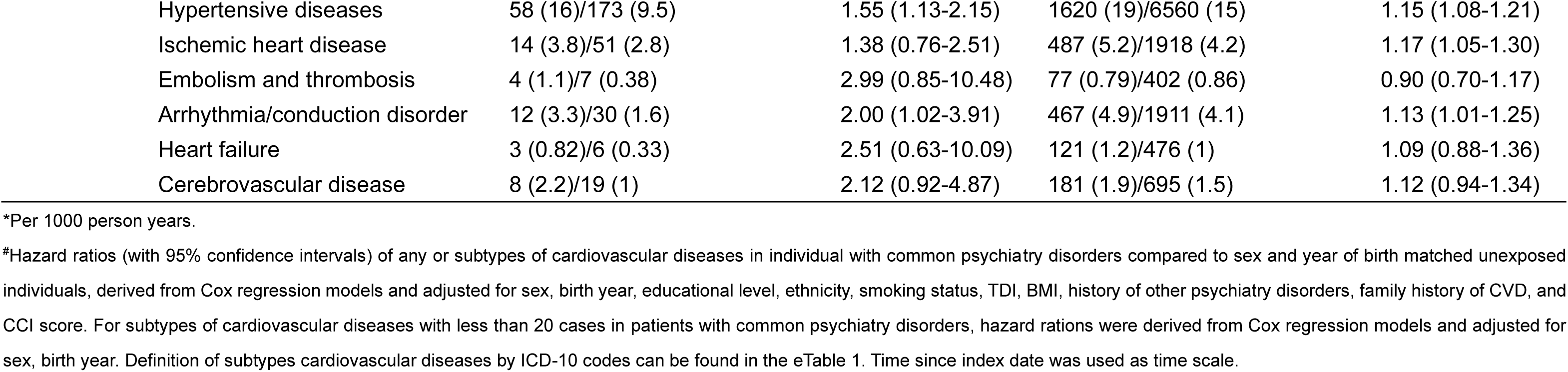
Hazard ratios (HRs) with 95% confidence intervals (CIs) of cardiovascular disease among patients with common psychiatry disorders compared to matched unexposed individuals, by different psychiatry disorders

**eTable 4.** PheWAS results of hazard ratios (HRs) with 95% confidence intervals (CIs) of different individual cardiovascular diseases among patients with common psychiatry disorders compared to matched unexposed individuals

| Cardiovascular diseases | Codes* | Case numbers<sup>#</sup> | HR (95% CIs)<sup>&</sup> | p |
| --- | --- | --- | --- | --- |
| Sequelae of cerebrovascular disease | I69 | 155 | 1.25 (1.02-1.54) | 0.0296 |
| Peripheral vascular disease | I73 | 198 | 1.29 (1.09-1.52) | 0.00307 |
| Chronic rheumatic heart disease | I05 | 275 | 0.84 (0.73-0.96) | 0.0106 |
| Embolism and thrombosis | I26 | 502 | 1.12 (1.01-1.24) | 0.037 |
| Other cerebrovascular diseases | I65 | 551 | 1.38 (1.24-1.53) | 1.77E-09 |
| Non-rheumatic valve disorders | I34 | 566 | 1.04 (0.94-1.15) | 0.411 |
| Complications and ill-defined descriptions of heart disease | I51 | 605 | 1.10 (1.00-1.21) | 0.041 |
| Heart failure | I50 | 637 | 1.08 (0.98-1.19) | 0.103 |
| Stroke | I60 | 694 | 1.20 (1.09-1.32) | 0.000129 |
| Acute myocardial infarction | I21 | 871 | 1.18 (1.09-1.29) | 0.000117 |
| Other cardiac arrhythmias | I49 | 1370 | 1.16 (1.09-1.24) | 7.83E-06 |
| Atrial fibrillation and flutter | I48 | 1404 | 1.03 (0.97-1.09) | 0.357 |
| Angina pectoris | I20 | 1521 | 1.30 (1.22-1.38) | 5.03E-17 |
| Chronic ischaemic heart disease | I25 | 2208 | 1.20 (1.14-1.26) | 5.96E-13 |
| Primary hypertension | I10 | 8239 | 1.16 (1.13-1.19) | 5.67E-28 |

\*Combined ICD-10 code; mapping between the original ICD-10 code and the combined ICD-10 code can be found in the eTable 2;
<sup>#</sup>Numbers of the individuals of common psychiatry disorders diagnosed with the corresponding cardiovascular diseases after the diagnosis of common psychiatry disorders;
<sup>&</sup>Hazard ratios (with 95% confidence intervals) of different cardiovascular diseases more than 150 cases in patients with common psychiatry disorders compared to sex and year of birth matched individuals without psychiatry disorders, derived from Cox regression models and adjusted for sex, birth year, educational level, ethnicity, smoking status, TDI, BMI, history of other psychiatry disorders, family history of CVD, and CCI score.

**eTable 5.** Odds ratios (ORs) with 95% confidence intervals (CIs) for the significant pairs of cardiovascular diseases following a diagnosis of common psychiatry disorders

| **Codes of CVD pairs\*** | **No.#** | **OR (95% CI)<sup>&</sup>** | **Percentage<sup>§</sup>** |
| --- | --- | --- | --- |
| I10-I20 | 505 | 5.01 (4.17-6.01) | 30.98% |
| I10-I21 | 224 | 4.02 (3.10-5.20) | 31.45% |
| I10-I25 | 739 | 4.60 (3.97-5.33) | 26.55% |
| I10-I49 | 419 | 2.97 (2.48-3.56) | 19.88% |
| I10-I60 | 203 | 3.81 (2.88-5.05) | 32.85% |
| I10-I65 | 226 | 5.10 (3.80-6.86) | 21.47% |
| I20-I49 | 115 | 3.12 (2.33-4.18) | 24.00% |
| I21-I20 | 164 | 24.89 (14.15-43.79) | 25.00% |
| I21-I25 | 193 | 29.32 (16.72-51.42) | 55.94% |
| I25-I20 | 430 | 21.65 (15.53-30.17) | 36.83% |
| I25-I49 | 174 | 4.46 (3.41-5.84) | 26.34% |
| I25-I65 | 79 | 2.84 (1.99-4.07) | 17.65% |

A total of 12 CVD pairs with N ≥ 75 and statistically significant odds ratios after Bonferroni correction for patients with common psychiatry disorders were show in this table;
Abbreviation: CVD, cardiovascular disease;
\*Combined ICD-10 code for two CVDs; mapping between the original ICD-10 code and the combined ICD-10 code can be found in the eTable 2;

**eTable 6.**
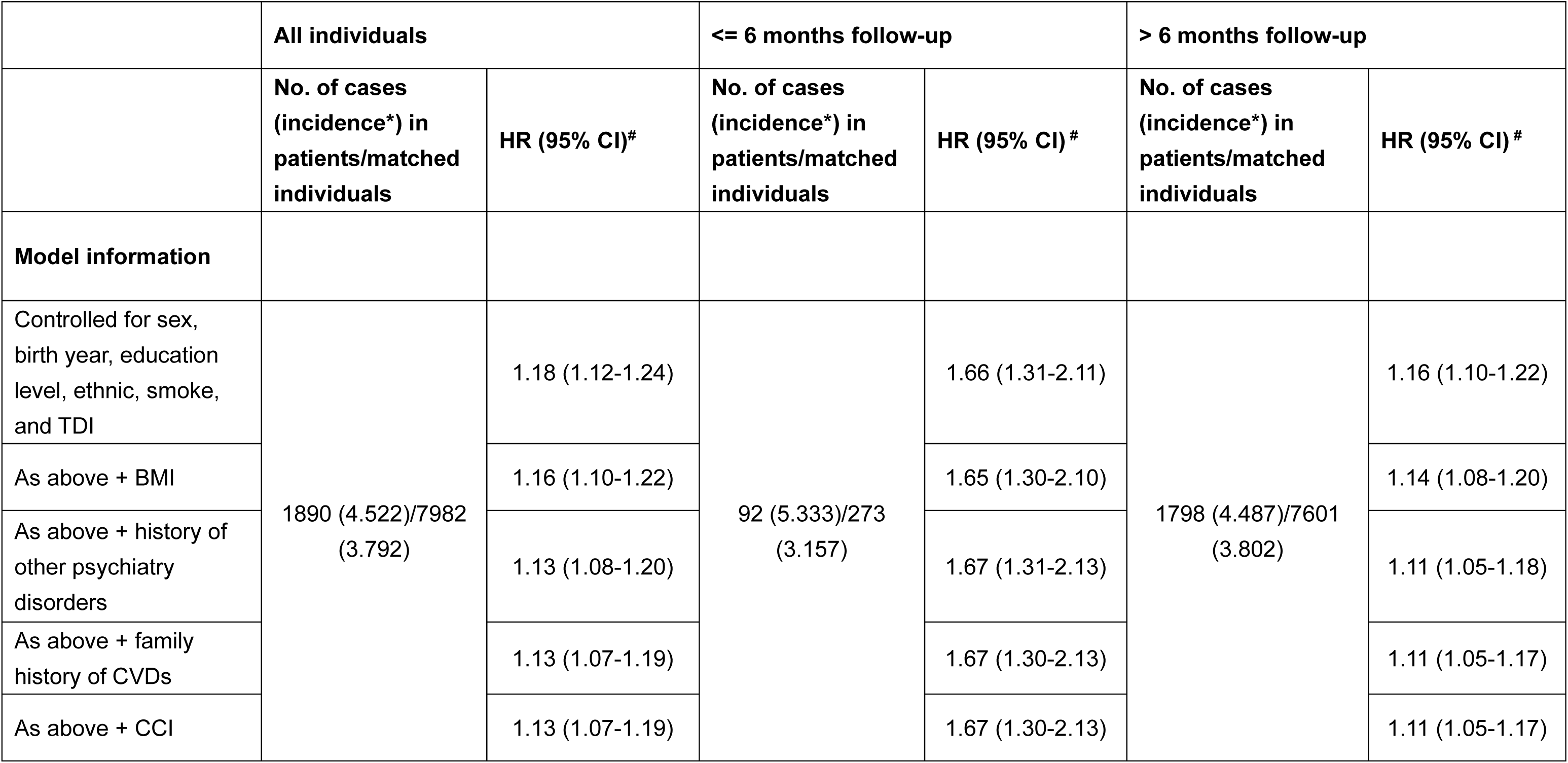
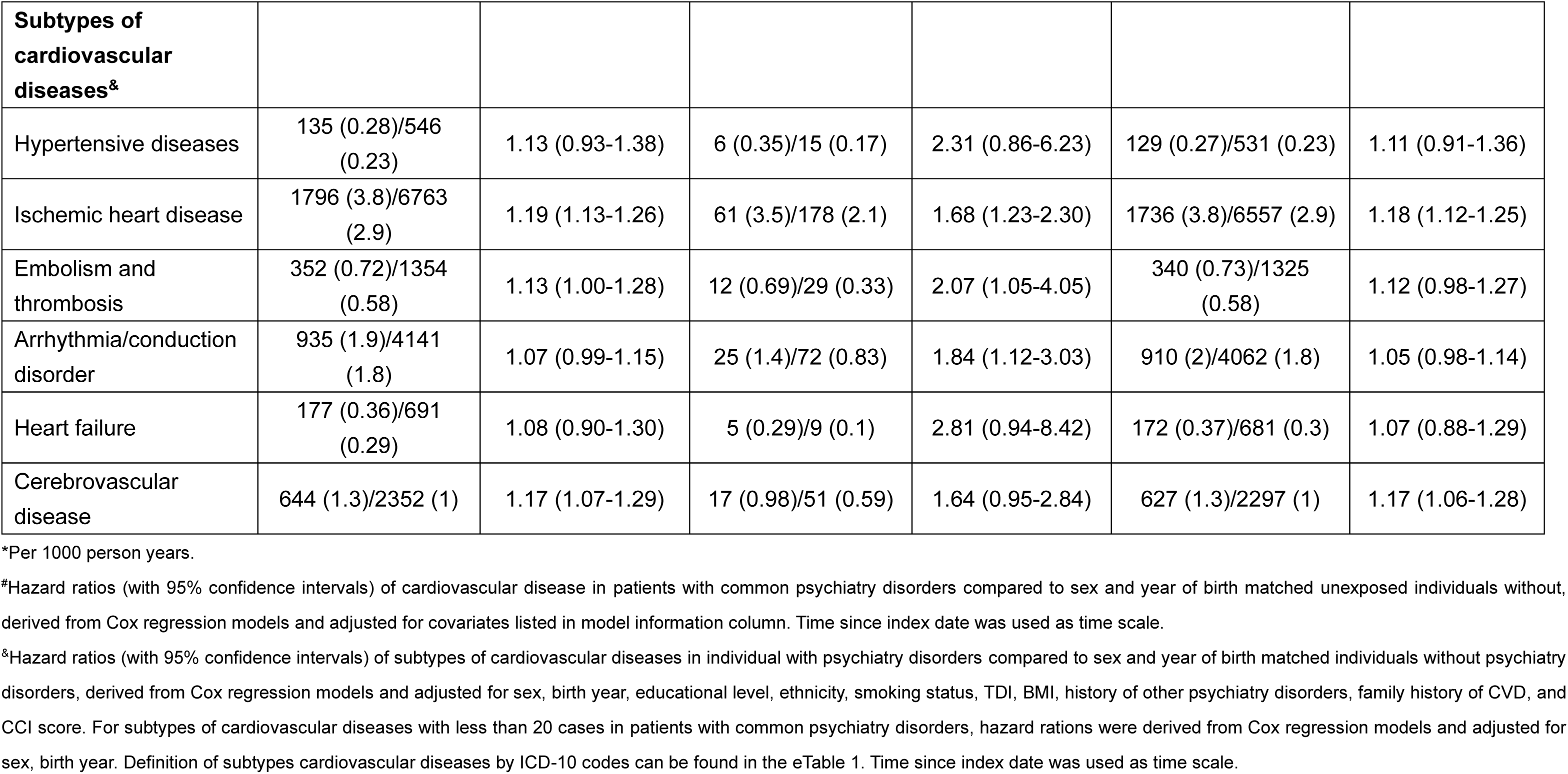
Hazard ratios (HRs) with 95% confidence intervals (CIs) of primary diagnosis of cardiovascular disease among patients with common psychiatry disorders compared to matched unexposed individuals

|  | All individuals |  | <= 6 months follow-up |  | > 6 months follow-up |  |
| --- | --- | --- | --- | --- | --- | --- |
|  | No. of cases<br>(incidence*) in<br>patients/matched<br>individuals | HR (95% CI)# | No. of cases<br>(incidence*) in<br>patients/matched<br>individuals | HR (95% CI) # | No. of cases<br>(incidence*) in<br>patients/matched<br>individuals | HR (95% CI) # |
| **Model information** |  |  |  |  |  |  |
| Controlled for sex,<br>birth year, education<br>level, ethnic, smoke,<br>and TDI | 1890 (4.522)/7982<br>(3.792) | 1.18 (1.12-1.24) | 92 (5.333)/273<br>(3.157) | 1.66 (1.31-2.11) | 1798 (4.487)/7601<br>(3.802) | 1.16 (1.10-1.22) |
| As above + BMI |  | 1.16 (1.10-1.22) |  | 1.65 (1.30-2.10) |  | 1.14 (1.08-1.20) |
| As above + history of<br>other psychiatry<br>disorders |  | 1.13 (1.08-1.20) |  | 1.67 (1.31-2.13) |  | 1.11 (1.05-1.18) |
| As above + family<br>history of CVDs |  | 1.13 (1.07-1.19) |  | 1.67 (1.30-2.13) |  | 1.11 (1.05-1.17) |
| As above + CCI |  | 1.13 (1.07-1.19) |  | 1.67 (1.30-2.13) |  | 1.11 (1.05-1.17) |

| **Subtypes of cardiovascular diseases<sup>&</sup>** |  |  |  |  |  |  |
| --- | --- | --- | --- | --- | --- | --- |
| Hypertensive diseases | 135 (0.28)/546 (0.23) | 1.13 (0.93-1.38) | 6 (0.35)/15 (0.17) | 2.31 (0.86-6.23) | 129 (0.27)/531 (0.23) | 1.11 (0.91-1.36) |
| Ischemic heart disease | 1796 (3.8)/6763 (2.9) | 1.19 (1.13-1.26) | 61 (3.5)/178 (2.1) | 1.68 (1.23-2.30) | 1736 (3.8)/6557 (2.9) | 1.18 (1.12-1.25) |
| Embolism and thrombosis | 352 (0.72)/1354 (0.58) | 1.13 (1.00-1.28) | 12 (0.69)/29 (0.33) | 2.07 (1.05-4.05) | 340 (0.73)/1325 (0.58) | 1.12 (0.98-1.27) |
| Arrhythmia/conduction disorder | 935 (1.9)/4141 (1.8) | 1.07 (0.99-1.15) | 25 (1.4)/72 (0.83) | 1.84 (1.12-3.03) | 910 (2)/4062 (1.8) | 1.05 (0.98-1.14) |
| Heart failure | 177 (0.36)/691 (0.29) | 1.08 (0.90-1.30) | 5 (0.29)/9 (0.1) | 2.81 (0.94-8.42) | 172 (0.37)/681 (0.3) | 1.07 (0.88-1.29) |
| Cerebrovascular disease | 644 (1.3)/2352 (1) | 1.17 (1.07-1.29) | 17 (0.98)/51 (0.59) | 1.64 (0.95-2.84) | 627 (1.3)/2297 (1) | 1.17 (1.06-1.28) |

\*Per 1000 person years.
<sup>#</sup>Hazard ratios (with 95% confidence intervals) of cardiovascular disease in patients with common psychiatry disorders compared to sex and year of birth matched unexposed individuals without, derived from Cox regression models and adjusted for covariates listed in model information column. Time since index date was used as time scale.
<sup>&</sup>Hazard ratios (with 95% confidence intervals) of subtypes of cardiovascular diseases in individual with psychiatry disorders compared to sex and year of birth matched individuals without psychiatry disorders, derived from Cox regression models and adjusted for sex, birth year, educational level, ethnicity, smoking status, TDI, BMI, history of other psychiatry disorders, family history of CVD, and CCI score. For subtypes of cardiovascular diseases with less than 20 cases in patients with common psychiatry disorders, hazard ratios were derived from Cox regression models and adjusted for sex, birth year. Definition of subtypes cardiovascular diseases by ICD-10 codes can be found in the eTable 1. Time since index date was used as time scale.

**eTable 7.** Hazard ratios (HRs) with 95% confidence intervals (CIs) of primary diagnosis of cardiovascular disease among patients with common psychiatry disorders compared to matched unexposed individuals, by different disease susceptibility

| Disease susceptibility | <= 6 months follow-up |  | > 6 months follow-up |  |
| --- | --- | --- | --- | --- |
|  | No. of cases (incidence*) in patients/matched individuals | HR (95% CI)<sup>#</sup> | No. of cases (incidence*) in patients/matched individuals | HR (95% CI)<sup>#</sup> |
| **Family history of cardiovascular disease** |  |  |  |  |
| No | 29 (3.951)/35 (2.041) | 1.88 (1.04-3.38) | 636 (3.668)/1087 (2.714) | 1.25 (1.12-1.40) |
| Yes | 63 (6.357)/105 (3.732) | 1.94 (1.38-2.72) | 1162 (5.112)/2963 (4.598) | 1.09 (1.01-1.17) |
| P<sub>interaction</sub> |  | 0.9283 |  | 0.0445 |
| **Level of polygenic risk score (PRS) of cardiovascular disease<sup>§</sup>** |  |  |  |  |
| Low (<the lower tertile) | 17 (3.756)/17 (2.943) | 1.35 (0.57-3.20) | 448 (4.19)/415 (3.06) | 1.48 (1.25-1.75) |
| Moderate (lower-upper tertile) | 31 (6.817)/20 (3.442) | 2.02 (1.14-3.56) | 440 (4.12)/525 (3.857) | 1.02 (0.87-1.19) |
| High (>the upper tertile) | 29 (6.3)/22 (3.72) | 1.93 (0.62-6.02) | 579 (5.453)/656 (4.782) | 1.19 (1.04-1.37) |
| P<sub>interaction</sub><sup>*</sup> |  | 0.6241 |  | 0.0493 |

\*Per 1000 person years.
<sup>#</sup>Cox regression models were stratified by disease susceptibility and adjusted for sex, birth year, educational level, ethnicity, smoking status, TDI, BMI, history of other psychiatry disorders, and CCI score.
<sup>§</sup>Polygenic risk score of cardiovascular disease were calculated based on published GWAS summary statistics data.
\* The differences in hazard ratios for CVD polygenic risk score were assessed between low and high subgroups by Wald test.

**eTable 8.** Odds ratios (ORs) with 95% confidence intervals (CIs) for the significant pairs of primary diagnosis of cardiovascular diseases following a diagnosis of common psychiatry disorders

| **Codes of CVD pairs\*\*** | **No.#** | **OR (95% CI)<sup>&</sup>** | **Percentage<sup>§</sup>** |
| --- | --- | --- | --- |
| I21-I20 | 89 | 19.77 (9.96-39.24) | 7.87% |
| I21-I25 | 184 | 29.99 (16.73-53.76) | 15.52% |

A total of 2 CVDs pairs with N ≥ 75 and statistically significant odds ratios after Bonferroni correction for individuals with psychiatry disorders were show in this table;
Abbreviation: CVD, cardiovascular disease;
\*Combined ICD-10 code for two CVDs; mapping between the original ICD-10 code and the combined ICD-10 code can be found in the eTable 2;

## Notes

### Competing Interest Statement

The authors have declared no competing interest.

